# Size matters: large copy number losses reveal novel Hirschsprung disease genes

**DOI:** 10.1101/2020.11.02.20221481

**Authors:** Laura Kuil, Katherine C. MacKenzie, Clara S Tang, Jonathan D. Windster, Thuy Linh Le, Anwarul Karim, Bianca M. de Graaf, Robert van der Helm, Yolande van Bever, Cornelius E.J. Sloots, Conny Meeussen, Dick Tibboel, Annelies de Klein, René M. H. Wijnen, Jeanne Amiel, Stanislas Lyonnet, Maria-Mercè Garcia-Barcelo, Paul K.H. Tam, Maria M. Alves, Alice Brooks, Robert M.W. Hofstra, Erwin Brosens

## Abstract

**Background:** Hirschsprung disease (HSCR) is characterized by absence of ganglia in the intestine. Approximately 18% of patients have additional anatomical malformations or neurological symptoms (HSCR-AAM). HSCR is a complex genetic disease in which the loss of enteric ganglia stems from a combination of genetic alterations: rare coding variants, predisposing haplotypes and Copy Number Variation (CNV). Pinpointing the responsible culprits within a large CNV is challenging as often many genes are affected. We investigated if we could find deleterious CNVs and if we could identify the genes responsible for the aganglionosis.

**Results:** Deleterious CNVs were detected in three groups of patients: HSCR-AAM, HSCR patients with a confirmed causal genetic variant and HSCR-isolated patients without a known causal variant and controls. Predisposing haplotypes were determined, confirming that every HSCR subgroup had increased contributions of predisposing haplotypes, but their contribution was highest in isolated HSCR patients without *RET* coding variants. CNV profiling proved that HSCR-AAM patients had larger copy number losses. Gene enrichment strategies using mouse enteric nervous system transcriptomes and constraint metrics were used to determine plausible candidate genes in Copy Number Losses. Validation in zebrafish using CRISPR/Cas9 targeting confirmed the contribution of *UFD1L, TBX2, SLC8A1* and *MAPK8* to ENS development. In addition, we revealed epistasis between reduced Ret and Gnl1 expression *in vivo*.

**Conclusion:** Rare large Copy Number losses - often *de novo* - contribute to the disease in HSCR-AAM patients specifically. We proved the involvement of five genes in enteric nervous system development and Hirschsprung disease.

## Introduction

Hirschsprung disease (HSCR) is characterized by absence of the enteric nervous system (ENS) in the intestine (aganglionosis). HSCR is a complex genetic disease where the phenotype stems from a combination of genetic alterations: rare coding variants, predisposing haplotypes and Copy Number Variation (CNV)[1]. Most patients have HSCR as an isolated congenital anomaly, while in others the absence of enteric neurons is part of a spectrum of anomalies of a monogenetic syndrome[2]. Approximately 18% of patients have additional anatomical malformations or neurological symptoms[3] (HSCR-AAM). Deletions of 10q11[4, 5] led to the identification of the REarranged during Transfection gene (*RET)*, the major responsible gene for familial and sporadic isolated HSCR[6, 7]. HSCR penetrance is influenced by predisposing *RET* risk haplotypes which increase disease risk substantially, especially if homozygous, in specific combinations (rs2506030, rs7069590, rs2435357)[8, 9], together with other risk loci near the semaphorin gene cluster (rs80227144)[9, 10], or the neuregulin 1 gene (*NRG1*; rs7005606)[11-13].

For many of HSCR-AAM patients the underlying cause is unknown. It is plausible that the associated anomalies in these patients are the result of a pathogenic single nucleotide variant or insertion/deletion. However, all different genomic variation types should be considered, since the presence of unbalanced translocations and CNV has resulted in the identification of causal genes for several syndromes with HSCR as a key feature. For example, deletions of 13q[1, 14-18] resulted in the identification of one of the genes responsible for Waardenburg-Shah syndrome type 4 (*EDNRB*)[19]. Deletions of 2q[20-23] and 4p[24] contributed to the discovery of genes responsible for Mowatt-Wilson syndrome (*ZEB2*, formerly *ZFHX1B*)[25] and the gene responsible for Congenital Central Hypoventilation Syndrome *(PHOX2B)*[26]. Furthermore, deletions (17q21[27] and 22q11[1, 28]) and duplications (17q23[29], dup22p[27, 30] and 22q11[31]) have been described in HSCR patients. In the light of these findings, we hypothesized that rare CNVs significantly contribute to aganglionosis in HSCR-AAM patients who lack a pathogenic coding variant in any of the known HSCR genes.

## Results

### Rare deletions are enriched in HSCR-AAM patients

Patient and control CNVs were determined and classified as a “rare CNV” when absent from large control cohorts (n=19,584). We also included known pathogenic or modifier CNVs[32, 33]. The size, type and gene content characteristics of rare CNVs were compared to those of 326 controls (Group 4, n=326). With this approach, 56 rare CNVs were detected in 34 HSCR patients (S3). We determined segregation of the rare CNVs in nine patients, five of these were *de novo*: three gains (22q11.21 - q11.22, 22q11.21 and 7q36.1) and two losses (17q23.1 - q23.2 and 6p22.1 - p21.33). CNV regions did not have a lot of overlap within our cohort, previously published CNV studies or with those described in the DECIPHER (https://decipher.sanger.ac.uk/) database (S3). The number of rare CNVs per individual did not differ between subgroups or controls (S4). However, CNVs found in HSCR-AAM patients (Group 1) were larger compared to controls (Group 4, p=7.297E-6, Figure 2A). This difference is attributed to outliers, large size of losses in specific patients of group 1 (p=3.15E-08)(S5), strongly suggesting a role of these specific CNVs in HSCR-AAM patients.

**Figure 1.**
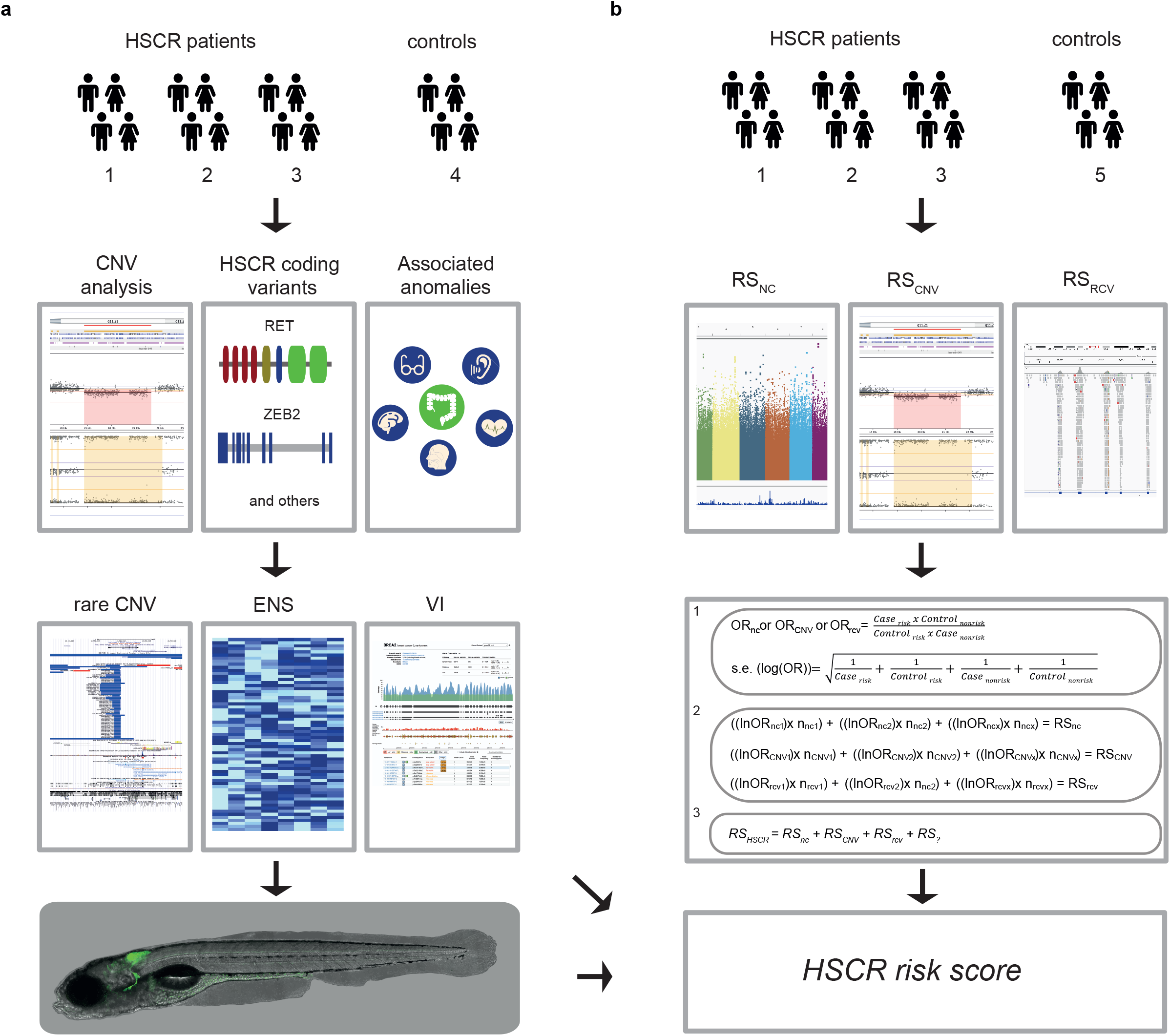
Schematic overview of our overall study design and methods used. a) In brief: We determined the Copy Number profiles of different subgroups of HSCR patients (1-3) and controls (4). We determined RET and / or known disease gene coding mutations and ranked the genes affected by a CNV according to frequency in controls, expression in the developing mouse ENS and gene tolerance to variation. Next, we determined whether disruption of the main candidate genes resulted in a reduction of enteric neurons in zebrafish. b) In parallel, we evaluated the contribution of different genetic risk factors, by comparing the contribution of known non-coding predisposing haplotypes across groups. o/e = overexpressed, VI = intolerant to variation.

**Figure 2.**
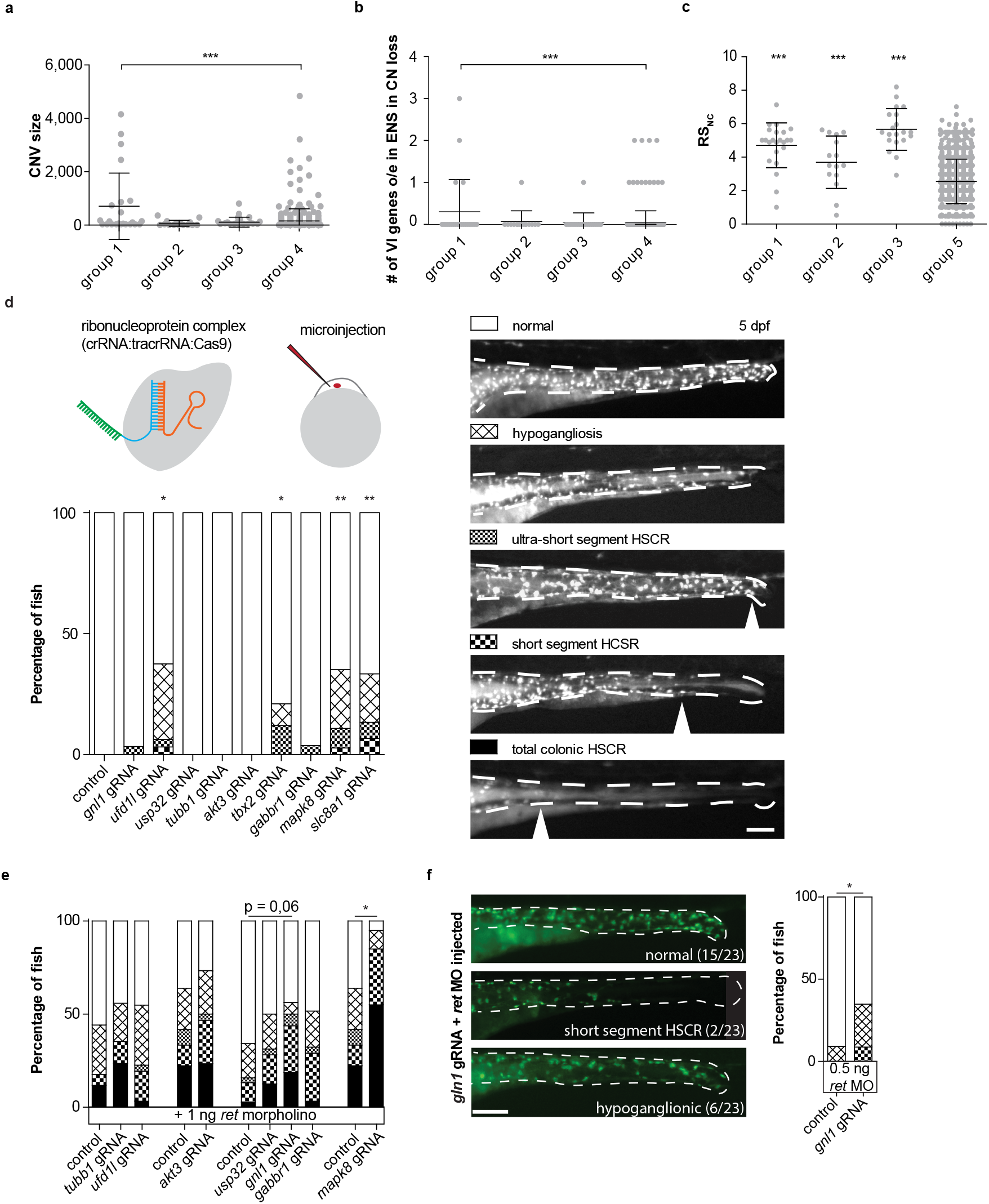
ENS overexpressed genes intolerant to variation were enriched in HSCR-complex patients and their disruption in zebrafish caused HSCR phenotypes. a) Graph showing CNV size, each dot represents a CNV. b) graph showing the number of variant intolerant (VI) genes overexpressed in the mouse ENS per patient, each dot represents one patient. c) Graph showing the RSnc, each dot represents one patient. Error bars represent standard deviation, statistical analysis used: students t-test (a-c). d) CRISPR/Cas9 complex injections in zebrafish fertilized eggs induced HSCR phenotypes upon disruption of four genes (accumulated data from multiple experiments). e) Graph showing accumulated data of the percentage of fish with HSCR phenotypes upon injection of a morpholino targeting Ret translation at a concentration that induced HSCR phenotypes in approximately 50% of the fish. Ret morpholino injections in combination with disruption of mapk8 shows epistasis. Disruption of gnl1 showed a trend towards higher penetrance of HSCR phenotypes. f) A second injection round targeting gnl1, using a lower dose of ret morpholino, confirmed gnl1 epistasis with Ret.

### Candidate genes within a loss are constraint and expressed during ENS development

HSCR disease genes (S6) are often constraint and expressed in the affected tissue, genes within a CNV region associated with HSCR should share these characteristics (Figure 1A). In Group 1, rare CNVs are enriched for ENS genes (p=4.565E-6, S7). Moreover, losses in HSCR-AAM patients (Group 1) contained more ENS genes that are constraint[34, 35], compared to losses found in controls (Group 4, Figure 2B, p=0,0004). Therefore, constraint genes within CN losses identified in group 1, and expressed in the ENS were prioritized as candidate genes for HSCR: *AKT3, GNL1, GABBR1, SLC8A1, MAPK8, UFD1L, TBX2, USP32* and *TUBB* (Table 1).

**Table 1.**
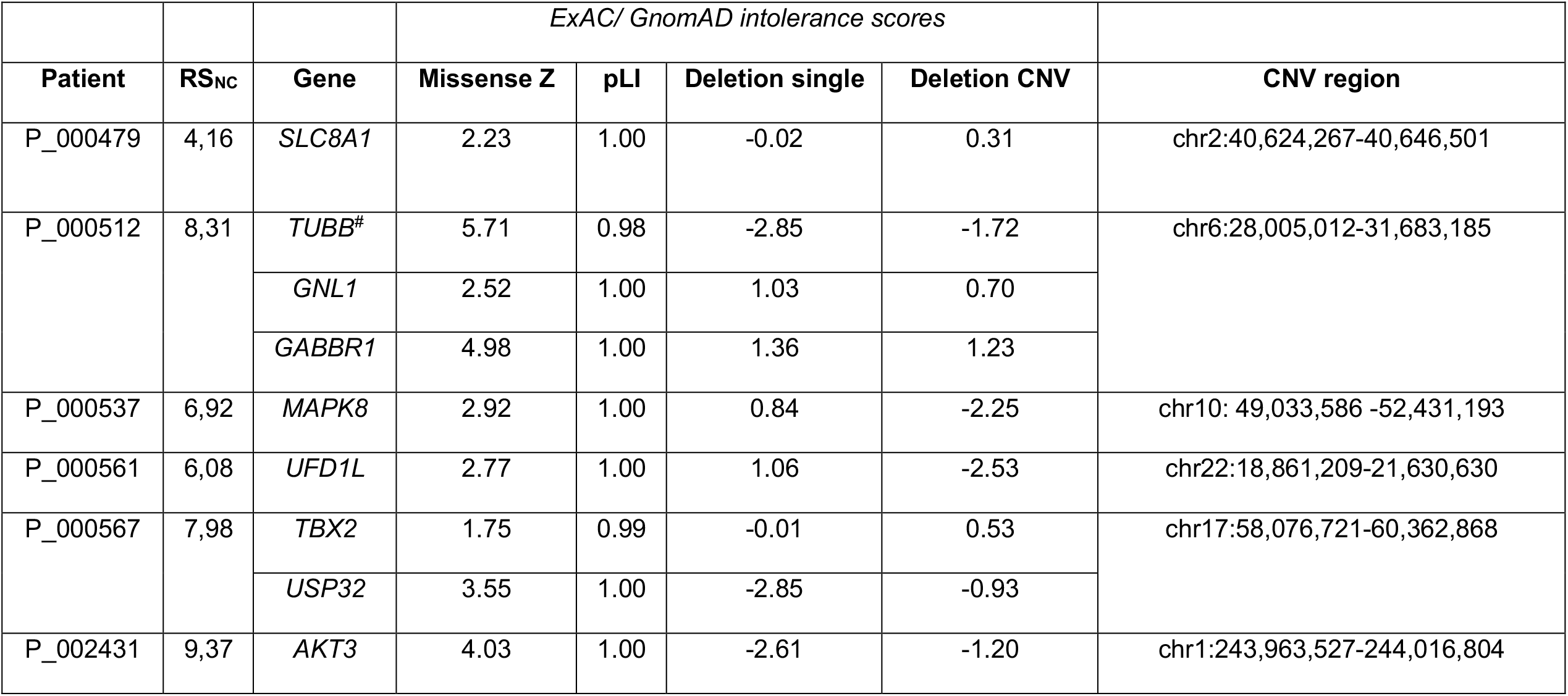
Genes in rare CN losses that are overexpressed in mouse ENS and intolerant to genetic variation. Genes marked with an # also have a loss of function variant in an independent HSCR cohort (see table 2). Depicted are the RSnc (see S11) and the CNV and variant intolerance scores derived from http://gnomad.broadinstitute.org/ and http://exac.broadinstitute.org/about.

### Zebrafish validation confirms the impact of candidate gene disruption on ENS development

To validate the effect of losing one copy of these constraint genes in ENS development and HSCR, we disrupted their orthologues in zebrafish by CRISPR/Cas9 targeting. Zebrafish are highly suited for reverse-genetic screening due to their rapid, *ex-utero* development and transparency and have proven instrumental in the identification of HSCR disease genes[36]. Premature termination of enteric neuron migration was observed in a subgroup of larvae injected with guide RNAs targeting *ufd1l* (p=0,0208), *tbx2* (p=0,0373), *slc8a1* (p=0,0073), and *mapk8* (p=0,0022, Figure 2D).

### HSCR disease coding genetics is heterogeneous

More evidence for their involvement could be provided by finding additional patients with putative deleterious alterations in genes affected by rare CNV. However, there was no significant enrichment for nonsense, splice site or missense variants in genes impacted by rare gains nor losses (S9) when comparing 443 short segment HSCR patients and 493 controls[37], underlining the genetic heterogeneity between HSCR patients. Nonsense or splice-site variants were detected in eight genes (Table 2). Only one variant, detected in DNA derived from blood and isolated ENS cells, affected a constraint gene overexpressed in the mouse ENS and present in a CN loss: a frameshift (*TUBB;* NM_001293212.1:c.1330_1331delCAinsA, p.Gln444Serfs*35) in a patient with isolated HSCR (table 1, S8).

**Table 2:** WES: Nonsense and splice site variants in rare CNV genes. Rare putative deleterious nonsense variants and variants predicted to affect splicing in a whole exome sequencing cohort of HSCR (n=76, 149 controls) [36] and whole genome sequencing cohort of 443 short segment HSCR patients and 493 unaffected controls[37]. Variants in genes intolerant to variation that were also impacted by the de novo 17q23.1 - q23.2 loss (INTS2, MED13), the de novo 6p22.1 - p21.33 loss (PRRC2A, TUBB), the 9p21 loss (LINGO2), the maternal inherited 10q11.22 - q11.23 loss (SGMS1), de novo 7q36.1 gain (KMT2C) and the 3q24 gain (SLC6A6).

| Sample | Chr | Start | Stop | Ref | Alt | Exon | Gene | Type | location | Effect | HGVS cDNA-level | CADD | gnomADv2.1 | gnomADv2.1 |
| --- | --- | --- | --- | --- | --- | --- | --- | --- | --- | --- | --- | --- | --- | --- |
|  |  |  |  |  |  |  |  |  |  |  |  |  | Exomes | Genomes |
| SE14-0527 | 17 | 59946466 | 59946466 | . | T | 23 | INTS2 | insertion | exonic | frameshift | NM_001330417.1:c.3172dupA | . | 0 | 0 |
| HK19-0006 | 17 | 60072727 | 60072727 | C | T | 10 | MED13 | snv | intronic | splicing | NM_005121:c.1968-1G>A | 22.4 | 0 | 0 |
| SE16-3114 | 17 | 59469360 | 59469360 | C | T | 26 | BCAS3 | snv | exonic | stopgain | NM_001320470.1:c.2773C>T | 18.4 | 0.0002 | 0.0007 |
| SE14-0656 | 6 | 31604286 | 31604286 | G | T | 27 | PRRC2A | snv | splicing | splicing | NM_004638.3:c.5836-1G>T | 23.6 | 0.0001 | 3.24E-05 |
| SE16-3123 | 6 | 30692109 | 30692110 | CA | A | 4 | TUBB | substitution | exonic | frameshift | NM_001293212.1:c.1330_1331delCAinsA | . | 0 | 0 |
| SE17-3220 | 7 | 151962296 | 151962296 | T | C | 8 | KMT2C | snv | splicing | splicing | NM_170606.2:c.1013-2A>G | 22 | 8.21E-06 | 0 |
| HK19-0002 | 9 | 28476025 | 28476025 | T | G |  | LINGO2 | snv | intronic | splicing | NM_001258282:c.-395-2A>C | 23.2 | 0 | 0 |
| HK19-0003 | 10 | 52103343 | 52103344 | TA | T | 7 | SGMS1 | deletion | exonic | frameshift | NM_147156:c.T529delT;p.F177del | 33 | 0 | 0 |
| HK19-0004 | 10 | 52104106 | 52104108 | CTG | C |  | SGMS1 | deletion | intronic | splicing | NM_147156:c.-313-2CAG>--G | 24.5 | 0 | 0 |
| HK19-0005 | 10 | 52349911 | 52349911 | A | G |  | SGMS1 | snv | intronic | splicing | NM_147156:c.-683+2T>C | 23 | 0 | 0 |
| SE16-3109 | 22 | 21065731 | 21065731 | G | A | 51 | PI4KA | snv | exonic | stopgain | NM_058004.3:c.5821C>T | 51 | 0.0002 | 0.0002 |
| HK19-0001 | 3 | 14485130 | 14485130 | A | G |  | SLC6A6 | snv | intronic | splicing | NM_001134367:c.297-6A>G | 15.92 | 0 | 0 |

### Copy number loss alone is likely not sufficient to result in HSCR

Considering that aganglionosis has a low prevalence in patients with 22q11[32, 33, 38] and 17q23[39, 40] deletions, CNVs alone are probably not sufficient to cause HSCR. HSCR penetrance is influenced by predisposing risk haplotypes [8-13]. Five non-coding risk haplotypes were tested in our three patient groups and compared to an *in-house* population dataset (group 5) (n=727, Risk non-coding (RSnc) =2.54). As expected, HSCR groups 1, 2 and 3 all had a higher RSnc compared to group 5 (p= 4.59825E-14, p= 0.000960603, p= 1.21875E-23, respectively) (Figure 2C), indicating a contribution of the risk haplotypes in all HSCR subgroups. (S10; Figure 1B). Associated common risk haplotypes have epistatic interactions, not only with each other, but also with known HSCR genes - such as *RET* and *NRG1*[10, 12, 41] - and modify HSCR penetrance in monogenetic syndromes and Down syndrome[42-46]. The effect of epistatic interactions is smaller in high penetrant HSCR monogenetic disorders and larger in disorders in which HSCR penetrance is lower[42, 43]. In line with this, contribution of these risk haplotypes was the lowest in group 2 (RSnc=3.70), followed by group 1 (RSnc=4.71), and the highest in group 3 (RSnc= 5.66). Thus, risk haplotypes have indeed a higher impact in HSCR patients without known coding variants, compared to those that contain known coding variants (S11). To model reduced *RET* expression in patients with predisposing haplotypes, we injected zebrafish with an antisense ATG blocking morpholino specific for *ret*. Disruption of the candidate genes *mapk8* (p=0,011, Figure 2E) and *gnl1* (p=0,0405, Figure 2F) in combination with the *ret* morpholino resulted in less neurons. Considering that disruption of *gnl1* alone did not induce a HSCR phenotype, these results suggest that only in a sensitized background loss of *gnl1* can increase HSCR penetrance.

### Candidate genes are associated to ENS development

CRISPR/CAS9 targeted knockdown of the candidate genes *SLC8A1, GNL1, MAPK8, UFD1L and TBX2* connects these genes to ENS development. These genes are overexpressed in mouse ENS and have associations to ENS development. For instance, Mitogen-Activated Protein Kinase 8 gene (*MAPK8*) is part of the Mitogen activated protein (MAP) kinase pathway and required for ENCCs to migrate properly[47]. *UFD1L*, as this encodes for a downstream target of HAND2[48]. *Hand2*^-/-^ mice have decreased numbers of enteric neurons, neuronal differentiation defects and a disorganized ENS[49, 50]. T-Box 2 (*TBX2*) codes for a transcription factor already known to be involved in the regulation of neural crest derived melanocytes[51], a process that is also hampered in specific HSCR patients carrying pathogenic variants in *EDN3, SOX10* and *EDNRB*[52, 53]. Candidate genes are discussed further in S8.

## Discussion

The results presented here highlight the importance of Copy Number profiling, leading us to propose a HSCR risk model that consists of risk scores for: rare coding variants, non-coding variants, rare CNVs and currently unknown risk factors (e.g. epigenetic modifications and perhaps environmental factors). Considering HSCR-AAM (group 1), 36% of patients have contained a CNV harboring genes overexpressed in the mouse ENS (Figure 3A). Our risk model (Figure 3C) indicates major contributing factors in each group: in group 1 CNVs, in group 2 coding variants and in group 3 noncoding variants (Figure 3A, C). The predisposing *RET* haplotypes reduce RET expression[54] and in combination with the other risk haplotypes increase HSCR risk substantially. Translating this population risk to a threshold for HSCR development in individual patients is challenging as it is not precisely known how many of (and if) these known predisposing haplotypes are sufficient to result in aganglionosis. If we set a threshold at a relatively high level (RSnc>6), this would explain HSCR in 30% of patients in group 3 (S11, Figure 2C, 3A). Most HSCR-AAM patients have a lower RSnc, which makes sense, as this group contains more high impact CNVs and whilst they lack *RET* coding variants, they potentially have variants in other genes (Figure 2C, 3A, S11). Translating our findings back to our full cohort including all patients seen by a clinical geneticist and screened for *RET* mutations, HSCR coding variants explain 21% of HSCR cases (Figure 3B). If we consider CNVs containing genes expressed in the ENS, we could explain an additional 10% (Figure 3B). Importantly, this is an underestimation, since for 55% of patients without a *RET* coding variant no copy number profiling is performed (Figure 3B) and we focussed exclusively on rare CNVs. Since more common CNVs can also modify HSCR penetrance[55, 56], our results emphasize the need for a more widespread genomic analyses in all subgroups. Noncoding risk scores could be one of the determinants for a more elaborate genetic evaluation. A high noncoding risk score in a HSCR patient without associated anomalies is less likely to have a deleterious RET coding variant, patients with a low score are more likely to benefit from exome sequencing. Similarly, patients with large *de novo* losses often have a more complex phenotype and more intensive clinical investigations might be indicated.

**Figure 3.**
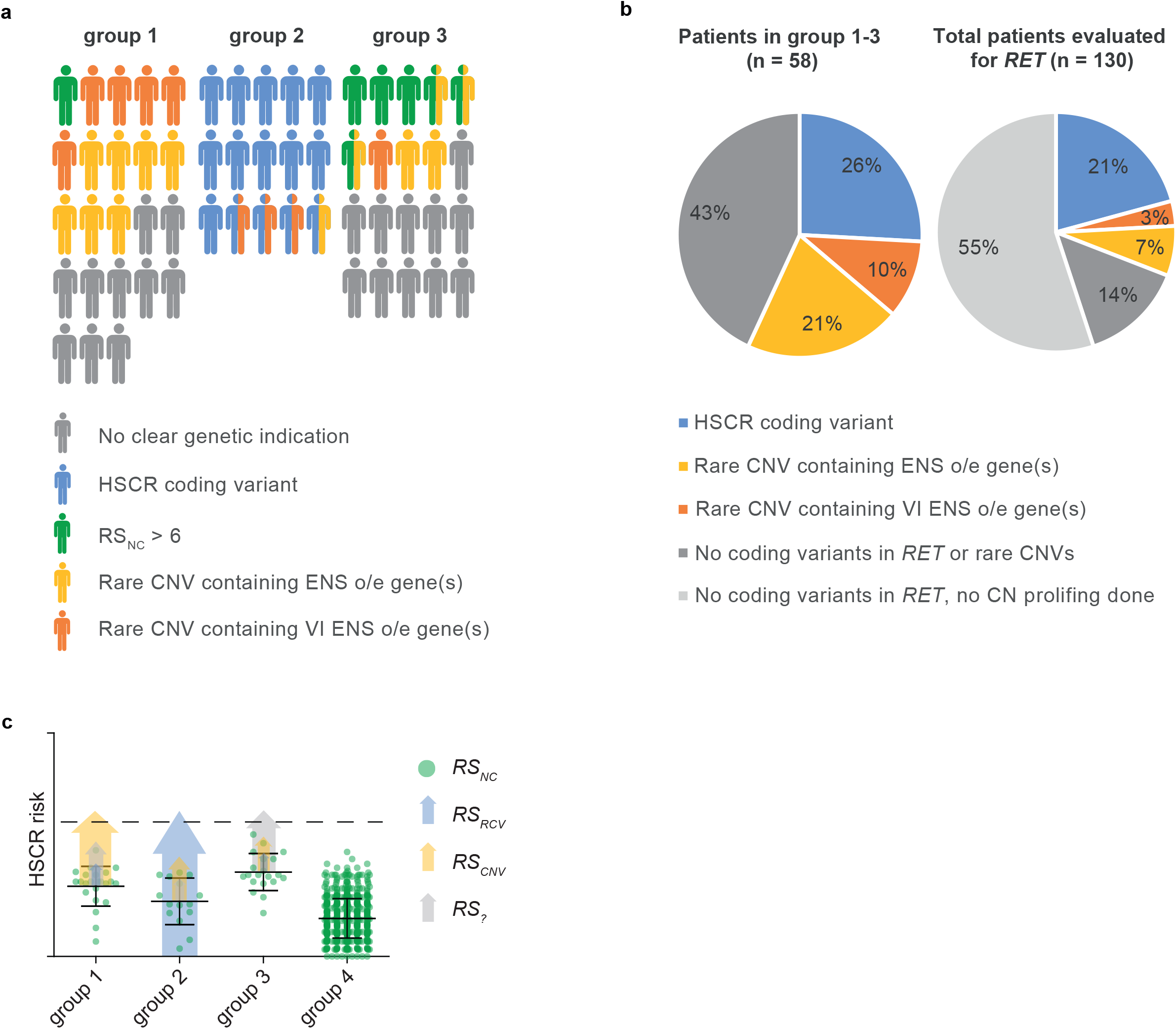
Complex HSCR genetics: genetic predispositions of HSCR patient groups. a) Visual representation of the distribution of genetic predispositions over HSCR patient groups. In total 197 patients born between 1973 and 2018 were evaluated by a clinical geneticist in the department of Clinical Genetics, Erasmus Medical Center, Rotterdam. Of these, 114 did not have associated anomalies nor a known syndrome. 29 patients had a known HSCR related genetic syndrome, including Down syndrome (n=18). 153 out of 197 patients were genetically evaluated for RET gene involvement and 21 had a pathogenic RET variant. b) Pie charts showing the incidence of rare CNVs containing genes overexpressed in the developing mouse ENS and coding variants in HSCR patients. c) Graphical representation of a hypothetical model explaining the relative contributions of the risk scores in our 3 patient groups. Error bars represent standard deviation.

## Conclusions

To summarize, HSCR genetics is complex with contributions of predisposing haplotypes in all HSCR subgroups. Rare large CNVs - often *de novo* (S5) - contribute substantially to the disease in HSCR-AAM patients. These CNVs are enriched for CN losses and for genes intolerant to variation that are overexpressed in the developing mouse ENS. Disruption of some of these genes in zebrafish confirmed that reduced expression of these genes increased the occurrence of HSCR, alone, or in epistasis with Ret. These genes have functional overlap with known HSCR disease genes: e.g. *UFD1L* is involved signalling receptor binding and *MAPK8* in axon guidance. Our “ENS expressed gene” based approach lead to the identification of new HSCR disease genes (Figure 3; Table 2): *UFD1L, TBX2, SLC8A1, GNL1* and *MAPK8*.

## Materials and methods

All authors had access to the study data and reviewed and approved the final manuscript

### Patient inclusion

This project was approved by the Medical ethics committee of the Erasmus Medical Centre (Hirschsprung disease: no 2012-582, addendum No. 1 and no.193.948/2000/159, addendum No. 1 and 2, MEC-20122387). We selected 58 out of 197 patients (Figure 3) for which DNA and informed consent were available, and in whom *RET* was screened. Three subgroups of HSCR patients were included in the CNV detection study: patients with HSCR and additional anatomical malformations or neurological defects, but without a *RET* pathogenic variant, or other causal genetic defect (group 1, n=23, S1), patients with HSCR and a known variant in *RET* or another causal gene (group 2, n=15, S2), and patients with only HSCR, without a deleterious *RET* coding variant or other causal genetic defect (group 3, n=20). Additionally, we included unaffected control individuals (group 4, n=326). Genotypes of noncoding predisposing loci were compared with those of other unaffected controls (group 5, n=727).

### Determination of Copy Number Variation (Figure 1a)

CNV profiles were determined with either the HumanCytoSNP-12 v2.1 or the Infinium Global Screening Array-24 v1.0 (Illumina Inc., San Diego, CA, USA), using methods previously described[57]. CNV profiles were inspected visually in Biodiscovery Nexus CN8.0 (Biodiscovery Inc., Hawthorne, CA, USA). CNVs with an overlap of at least 75% with similar state CN changes, were either classified as rare, when absent from large control cohorts (n=19,584), or as a known modifier[32, 33]. CNV number, size, type and gene content of rare CNVs were determined in HSCR patients (n=58) and unaffected controls (n=326) and compared between the control groups and previously described HSCR subgroups. All rare CNVs were uploaded to the ClinVar database (https://www.ncbi.nlm.nih.gov/clinvar/) and are depicted in S3.

### Exclusion of the involvement of known disease genes (Figure 1a)

The presence of *RET* coding mutations and those in intron-exon boundaries, in all patients in this study are determined. Furthermore, if a specific monogenetic syndrome (S6) was suspected based on the phenotypic spectrum observed, the suspected gene(s) were evaluated using a targeted NGS panel or whole exome sequencing. In four HSCR patients with associated anomalies (Group 1) and nine HSCR patients without associated anomalies (Group 3), the involvement of other known disease genes was excluded[2, 36, 37, 58] using whole exome sequencing (WES) with previously described pipelines[59, 60] and variant prioritization methods[61].

### Evaluation of candidate gene expression (Figure 1a)

We prioritized candidate genes based on gene characteristics: genes that are intolerant to variation and/or dosage sensitive (see variant filter criteria), and overexpressed in the developing mouse ENS compared to whole gut, between embryonic day E11.5 and E15.5 [62, 63]. Since data from human colon was only available for embryonic week 12, 14 and 16, we evaluated gene expression in these time points[64]. Data was downloaded from the gene expression omnibus (GSE34208 and GSE100130). Genes that were dosage sensitive, and either overexpressed in the mouse ENS or highly expressed in human embryonic colon, were selected.

### Variant prioritization in loss of function genes (Figure 1a)

To determine whether a gene affected by a rare putative deleterious CNV was constraint[34, 35], we allowed some tolerance to account for reduced penetrance (S9). Variants from NGS data previously generated were prioritized according to settings described in S9: These data included a WES cohort of sporadic HSCR (n=76, 149 controls) and (2) a Whole Genome Sequencing (WGS) cohort of 443 short segment HSCR patients and 493 unaffected controls[37]. Using RVTESTS[65], a variant burden test was done comparing the variant burden in 443 short segment HSCR patients and 493 controls (S9). All rare putative deleterious loss of function variants unique to the HSCR cohort in constraint genes are described in table 1.

### Genotyping of HSCR associated SNPs

Sanger sequencing was used to genotype all patients for SNPs known to be associated to HSCR[8-10, 55]. Primer sequences can be found in S11. We used (proxy) SNPs present on the GSAMD-v1 chip to determine the Rotterdam population background for these risk haplotypes (S10).

### Experimental animals

Zebrafish were kept on a 14h/10h light–dark cycle at 28°C, during development and adulthood. Tg(*phox2bb:*GFP) animals were used for all experiments[66]. Larvae were kept in HEPES-buffered E3 medium and 0.003% 1-phenyl 2-thiourea (PTU) was added 24 hours post fertilization (hpf), to prevent pigmentation.

### CRISPR/Cas9 gene disruption

Gene targeting using CRISPR/Cas9 was performed as described previously[67]. The Alt-R CRISPR-Cas9 System from Intergrated DNA Technologies (IDT) was used[68] (Figure 2A). crRNAs were designed using the IDT tool and selected on high target efficiency and low off target effect. gRNA sequences are listed in S13, primers used are listed in supplementary S14. Efficiency of indel generation was determined as described previously[67] (S15).

### Morpholino injections

Fertilized zebrafish eggs were injected with the CRISPR/Cas9 complex. Subsequently, 0,5 or 1 ng of translation blocking morpholino against *ret* (5′-ACACGATTCCCCGCGTACTTCCCAT-3′), and 1:10 Phenol Red (Sigma-Aldrich), was injected between the 1- and 4-cell stage (see figure 2D)[69]. To minimize variability, the same needle was used to inject the controls, that were not injected with CRISPR/Cas9 complex.

### Imaging

Images of 5 dpf tg(*phox2bb:*GFP) larvae were taken using a Leica M165 fluorescent dissection microscope with the Leica LASX software. Larvae were anesthetized with 0.016% MS-222 in HEPES buffered E3 medium and positioned on their lateral side on a 1.5 - 2% agarose coated petridish, to enable visualization of the enteric neurons.

### Scoring HSCR phenotypes in zebrafish

To determine whether the larvae present with a HSCR phenotype five categories were made: normal, hypoganglionic (less neurons), ultra-short segment HSCR (only most distal end of the gut lack neurons), short segment HSCR (neurons colonize until the midgut), and total colonic HSCR (neurons are absent from the total intestine) (see figure 2C).

### Statistical analysis

The number and size of rare CNVs, the number of rare losses and gains, the number of genes intolerant to variation (SNVs and CNVs), the number of genes overexpressed in mouse ENS per rare CNV, and the relative weighted risk score, were determined and compared for the different groups with a single ANOVA test. If group differences existed (P<0.05), we determined which subgroups were significantly different, using a two-tailed T-test. For statistical analysis of the zebrafish experiments the online tool MedCalc (MedCalc software ltd., Ostend, Belgium), was used together with the “N-1” Chi-squared test, number of larvae used and p values can be found in supplementary S16.

## Data Availability

All processed data generated or analyzed during this study is included in this manuscript and its supplementary information files. Any additional information or data of the current study are available from the corresponding author on reasonable request. All rare CNV are submitted to the ClinVar database (https://www.ncbi.nlm.nih.gov/clinvar/, SUB7813228).

## Supplemental Data description

CNV_HSCR_MedRxiv_Kuil_etal_supplement contains 16 elements including detailed patients descriptions, additional results, extended methods, primers and gRNA sequences.

## Author Contributions

Guarantor of the article [EB]; Conception or design of the work [EB, AB, RH], the acquisition [RW, JW, TL, RH, YB, CS, CM, DT, LK], analysis [LK, KM, CT, BM, AK], or interpretation [EB, LK, CT] of data; drafted the work [EB, KM, LK] or substantively revised it [MA, RH, AB, JA, SL, MB, PT].

## Acknowledgements

This study makes use of data generated by the DECIPHER community[70]. A full list of centers who contributed to the generation of the data is available from http://decipher.sanger.ac.uk and via email from.

