## supplementary S for "Size matters: large copy number losses reveal novel Hirschsprung disease genes"

**Index**

### S1: Hirschsprung patients without a RET mutation and additional phenotypical features

| Patient | **HSCR type** | **Other phenotypical characteristics** |
| --- | --- | --- |
| P_000482 | Short | Hydrocephalus, macrocephaly, autism |
| P_000540 | Short | Facial dysmorphisms |
| P_000494 | Short | Cardiac defects (VSD, ASD PDA, tricuspid atresia), dysplastic ears, renal malrotation |
| P_000512 | Short | Epilepsy, intellectual disability |
| P_000553 | Short | Cardiac defects (VSD, dextrocardia, PDA, double outlet right ventricle), intestinal malrotation |
| P_000559 | Total colonic | Dysmorphic features, tracheomalacia, cardiac defects (dilated left ventricle, absence of AV conduction) |
| P_000561 | Short | Facial dysmorphisms, small fontanelle, gastro-esophageal reflux, laryngeal web |
| P_000555 | Short | Hypoplastic thumb, hearing loss, developmental delay, facial dysmorphisms |
| P_002459 | Short | Hypospadias, mild autism |
| P_000567 | Short | Facial dysmorphisms, hearing loss, microcephaly, immunological hypersensitivity, nevus flammeus |
| P_000536 | Abnormal | Telecanthus upslant; short segment HSCR although a longer segment is abnormal ganglionated. |
| P_000562 | Short | Cafe au lait spots, cardiac defect (VSD) |
| P_000572 | TIA | Retrognathia, skin abnormality, facial dysmorphisms, cardiac defect (pulmonary valve stenosis) |
| P_000568 | Short | Dysmorphic features, hydrocele testis, hemangioma |
| P_000478 | Short | Hypertelorism, facial dysmorphisms |
| P_000520 | Short | Mild facial dysmorphisms, sandal-gap of toe |
| P_002455 | Short | Hypermobility of fingers; mild developmental delay, downslant of eyes |
| P_001763 | Short | White hair lock, mild developmental day |
| P_000537 | Short | Gross motor delay, spastic hemiplegia, bronchopulmonary dysplasia, cardiac defect (PDA) |
| P_000528 | Total colonic | Intellectual disability |
| P_000573 | Short | Epicanthal folds, small ears, broad eyebrows with mild synophrys |
| P_002450 | Long | Developmental delay |
| P_002343 | Short | Hypertelorism, long deeply grooved philtrum |

Depicted the patients with HSCR and additional anatomical malformations or neurological defects, but without a *RET* pathogenic variant, or other causal genetic defect (group 1, n=23)

### S2 HSCR patients with a deleterious variant

| Patient | HSCR type | Other phenotypical characteristics | Genetic defect |
| --- | --- | --- | --- |
| P_000302 | Short | no | NM_002181.3 (IHH):c.151C>A, NM_000168.5 (GLI3):c.2119C>T, NM_001190468.1 (GDNF):c.676_681delGGATGT |
| P_000526 | Short | no | NM_020630 (RET):c.1196C>T |
| P_000479 | Long | no | NM_020630 (RET):c.656-21C>T |
| P_000502 | Short | no | NM_020630 (RET):c.1880_1892del |
| P_002442 | Long | no | NM_020630 (RET):c.2599C>T |
| P_000566 | Short | no | NM_020630 (RET):c.3173A>G |
| P_000544 | Long | no | NM_020630 (RET):c.2690G>A |
| P_000534 | Short | hypospadias, anorectal malformation type perineal fistula | NM_020630 (RET):c.2906G>A |
| P_000480 | Short | Short stature | NM_013956 (NRG1):c.811A>T |
| P_004502 | Short | Hypertelorism, triangular face, pointy chin, straight eye brows, deepset eyes, small dysmorphic ears, agenesis of corpus callosum, hypospadia, dysmorphic nose | NM_014795 (ZEB2): c.1570del |
| P_000557 | Total colonic | postaxial polydactyly | NM_020630 (RET):c.C229C>T |
| P_000518 | Short | Facial dysmorphisms, microcephaly, bilateral generalized polymicogyria, developmental delay, short stature, hypotonia, eye anomaly | NM_015634 (KIFBP):c.268C>T |
| P_000486 | Total colonic | No abnormal phenotypedescribed. Normal psychomotor development | NM_001122659 (EDNRB):c.534_535insGGTGCCT |
| P_000570 | Short | congenital central hypoventilation syndrome | NM_003924 (PHOX2B):c.738_761dup |
| P_000576 | Short | Microcephaly, epicantus folds, upslant eyes, broad nose, synophrys naevus sacralis hyperpigmentosis back and shoulders, abnormal palmar creases | NM_020630 (RET):c.1321A>C, : NM_020630 (RET):c.C1941C>T |

Depicted the patients with HSCR and a known variant in *RET* or another causal gene (group 2, n=15)

### S3: rare CNV detected in this cohort

| ID | Chromosome Region | Length | Cytoband | Probes | Classification | Inheritance | Gender | Group | ClinVar ID |
| --- | --- | --- | --- | --- | --- | --- | --- | --- | --- |
| P_000479 | NC_000001.10:g.3776519_4049451dup | 272933 | p36.32 | 57 | VUS | undertermined | M | 2 | SCV001426225 |
| P_000544 | NC_000001.10:g.25715675_25744764dup | 29090 | p36.11 | 26 | VUS | undertermined | M | 2 | SCV001426226 |
| P_001636 | NC_000001.10:g.25715675_25744764dup | 29090 | p36.11 | 26 | VUS | undertermined | M | 3 | SCV001426226 |
| P_000566 | NC_000001.10:g.25717366_25744764dup | 27399 | p36.11 | 24 | VUS | undertermined | M | 2 | SCV001426227 |
| P_000498 | NC_000001.10:g.152286216_152323703dup | 37488 | q21.3 | 11 | VUS | undertermined | F | 3 | SCV001426228 |
| P_000450 | NC_000001.10:g.185109784_185132629dup | 22846 | q25.3 | 41 | VUS | undertermined | M | 3 | SCV001426229 |
| P_002431 | NC_000001.10:g.243963527_244016804del | 53278 | q44 | 9 | likely deleterious | undertermined | F | 3 | SCV001426230 |
| P_001637 | NC_000002.11:g.10664398_10914786dup | 250389 | p25.1 | 70 | VUS | undertermined | M | 3 | SCV001426231 |
| P_000479 | NC_000002.11:g.40624267_40646501del | 22235 | p22.1 | 11 | likely deleterious | undertermined | M | 2 | SCV001426232 |
| P_000582 | NC_000002.11:g.102658576_102847088dup | 188513 | q11.2 - q12.1 | 57 | VUS | undertermined | M | 3 | SCV001426233 |
| P_000567 | NC_000002.11:g.177128475_177259979dup | 131505 | q31.1 | 15 | VUS | undertermined | M | 1 | SCV001426234 |
| P_000557 | NC_000002.11:g.189848997_189872988del | 23992 | q32.2 | 53 | VUS | undertermined | M | 2 | SCV001426235 |
| P_000557 | NC_000002.11:g.206875802_207000559del | 124758 | q33.3 | 18 | VUS | undertermined | M | 2 | SCV001426236 |
| P_000573 | NC_000002.11:g.216214577_216299733del | 85157 | q35 | 49 | VUS | undertermined | F | 1 | SCV001426237 |
| P_000490 | NC_000002.11:g.220283220_220284269del | 1050 | q35 | 9 | VUS | undertermined | F | 3 | SCV001426238 |
| P_000302 | NC_000003.11:g.14406477_14509088dup | 102612 | p25.1 | 53 | likely deleterious | undertermined | F | 2 | SCV001426239 |
| P_000515 | NC_000003.11:g.57414966_57492375del | 77410 | p14.3 | 15 | VUS | undertermined | M | 3 | SCV001426240 |
| P_000579 | NC_000003.11:g.60468409_60490104del | 21696 | p14.2 | 16 | likely deleterious | undertermined | M | 3 | SCV001426241 |
| P_000512 | NC_000003.11:g.108481092_108926543del | 445452 | q13.13 | 30 | VUS | maternal | F | 1 | SCV001426242 |
| P_000480 | NC_000003.11:g.137727844_137778355del | 50512 | q22.3 | 14 | VUS | undertermined | M | 2 | SCV001426243 |
| P_002431 | NC_000003.11:g.145774557_145801149dup | 26593 | q24 | 9 | VUS | undertermined | F | 3 | SCV001426244 |
| P_000557 | NC_000004.11:g.159541187_159631380del | 90194 | q32.1 | 48 | VUS | undertermined | M | 2 | SCV001426245 |
| P_000515 | NC_000004.11:g.159596032_159621356del | 25325 | q32.1 | 20 | VUS | undertermined | M | 3 | SCV001426246 |
| P_001639 | NC_000006.11:g.22008230_22093109del | 84880 | p22.3 | 32 | VUS | undertermined | M | 3 | SCV001426247 |
| P_000512 | NC_000006.11:g.28005012_31683185del | 3678174 | p22.1 - p21.33 | 403 | likely deleterious | de novo | F | 1 | SCV001426248 |
| P_000490 | NC_000006.11:g.49662135_49664605del | 2471 | p12.3 | 6 | VUS | undertermined | F | 3 | SCV001426249 |
| P_002455 | NC_000007.13:g.3627221_3759274del | 132054 | p22.2 | 32 | VUS | undertermined | M | 1 | SCV001426250 |
| P_000582 | NC_000007.13:g.4736454_4860123dup | 123671 | p22.1 | 47 | VUS | undertermined | M | 3 | SCV001426251 |
| P_000582 | NC_000007.13:g.4929022_5218030dup | 289009 | p22.1 | 59 | VUS | undertermined | M | 3 | SCV001426252 |
| P_000582 | NC_000007.13:g.5239584_5401976dup | 162393 | p22.1 | 54 | VUS | undertermined | M | 3 | SCV001426253 |
| P_002431 | NC_000007.13:g.95845896_96004178del | 158283 | q21.3 | 16 | VUS | undertermined | F | 3 | SCV001426254 |
| P_000490 | NC_000007.13:g.117233848_117237342del | 3495 | q31.2 | 13 | VUS | undertermined | F | 3 | SCV001426255 |
| P_000490 | NC_000007.13:g.117287774_117293718del | 5945 | q31.2 | 6 | VUS | undertermined | F | 3 | SCV001426256 |
| P_000555 | NC_000007.13:g.151797921_152258693dup | 460773 | q36.1 | 52 | VUS | de novo | F | 1 | SCV001426257 |

### S3: rare CNV detected in this cohort (continued)

| ID | Chromosome Region | Length | Cytoband | Probes | Classification | Inheritance | Gender | Group | ClinVar ID |
| --- | --- | --- | --- | --- | --- | --- | --- | --- | --- |
| P_000568 | NC_000008.10:g.95186036_95301703dup | 115668 | q22.1 | 41 | VUS | undertermined | M | 1 | SCV001426258 |
| P_002450 | NC_000009.11:g.28393380_28462962del | 69583 | p21.1 | 21 | VUS | undertermined | M | 1 | SCV001426259 |
| P_000537 | NC_000010.10:g.49033586_52417694del | 3384109 | q11.22 - q11.23 | 183 | likely deleterious | maternal | M | 1 | SCV001426260 |
| P_000557 | NC_000011.9:g.62251301_62298871dup | 47571 | q12.3 | 25 | likely deleterious | undertermined | M | 2 | SCV001426261 |
| P_000479 | NC_000012.11:g.9245492_9308543dup | 63052 | p13.31 | 24 | VUS | undertermined | M | 2 | SCV001426262 |
| P_000561 | NC_000012.11:g.80598919_80836812del | 237894 | q21.31 | 12 | VUS | undertermined | M | 1 | SCV001426263 |
| P_002459 | NC_000012.11:g.94767704_94880489del | 112786 | q22 | 27 | VUS | undertermined | M | 1 | SCV001426264 |
| P_000494 | NC_000012.11:g.128208742_128917555dup | 708814 | q24.32 | 96 | VUS | paternal | M | 1 | SCV001426265 |
| P_000544 | NC_000015.9:g.62409198_62521004dup | 111807 | q22.2 | 33 | VUS | undertermined | M | 2 | SCV001426266 |
| P_000502 | NC_000015.9:g.80527215_80603142dup | 75928 | q25.1 | 22 | VUS | undertermined | F | 2 | SCV001426267 |
| P_000567 | NC_000017.10:g.58076721_60362868del | 2286148 | q23.1 - q23.2 | 74 | likely deleterious | de novo | M | 1 | SCV001426268 |
| P_000552 | NC_000018.9:g.6335542_6430944dup | 95403 | p11.31 | 20 | VUS | undertermined | M | 3 | SCV001426269 |
| P_000552 | NC_000018.9:g.6454093_6548624dup | 94532 | p11.31 | 37 | VUS | undertermined | M | 3 | SCV001426270 |
| P_000520 | NC_000018.9:g.45755986_45787673dup | 31688 | q21.1 | 13 | VUS | undertermined | M | 1 | SCV001426271 |
| P_001763 | NC_000018.9:g.65699090_66534856dup | 835767 | q22.1 | 228 | VUS | undertermined | F | 1 | SCV001426272 |
| P_000514 | NC_000021.8:g.47819478_47844620del | 25143 | q22.3 | 14 | VUS | undertermined | M | 3 | SCV001426273 |
| P_001632 | NC_000022.10:g.18687210_19060954dup | 373745 | q11.21 | 75 | VUS; modifier | undertermined | M | 3 | SCV001426274 |
| P_000561 | NC_000022.10:g.18861209_21630630del | 2769422 | q11.21 | 446 | likely deleterious | de novo | M | 1 | SCV001426275 |
| P_002455 | NC_000022.10:g.21802791_22555544dup | 752756 | q11.21 - q11.22 | 169 | VUS | de novo | M | 1 | SCV001426276 |
| P_000573 | NC_000023.10:g.3184901_3240953dup | 56053 | p22.33 | 16 | VUS | undertermined | F | 1 | SCV001426277 |
| P_000579 | NC_000023.10:g.94409037_94446394del | 37358 | q21.33 | 5 | VUS | undertermined | M | 3 | SCV001426278 |
| P_000540 | NC_000023.10:g.154277428_154299482del | 22055 | q28 | 5 | VUS | maternal | M | 1 | SCV001426279 |

**S3: rare CNV detected in this cohort (continued)**

In total, 56 rare CNVs were detected in HSCR patients. In group 1, 10 CN losses, 8 CN gains and 1 maternally inherited hemizygous loss on chromosome X in a male patient, were detected. In group 2, 5 CN losses and 8 CN gains were identified, and in group 3, 7 CN losses, 12 CN Gains, 4 homozygous losses and 1 hemizygous loss on chromosome X in a male patient, were found. We could determin segregation of the rare CNVs in nine patients, five of these were *de novo*. The inheritance pattern of other rare CNVs could not be determined due to unavailability of parental DNA. Although we did not determine segregation of all rare CNVs, this already suggests a high frequency of *de novo* CNVs in this cohort. Three gains (22q11.21 - q11.22, 22q11.21 and 7q36.1) and two losses (17q23.1 - q23.2 and 6p22.1 - p21.33), were *de novo*. Two CNVs were inherited maternally (10q11.22 - q11.23 loss and Xq28 loss). Two of the homozygous losses in patient P_000490 (group 3), disrupt the Cystic fibrosis transmembrane conductance regulator (*CFTR*) locus. However, this patient does not show signs of cystic fibrosis.

#### Overlap with previously described CNV and candidate genes in rare losses

Two regions overlapped in our small cohort: a gain in patients P_000544, P_000566 (group 2) and P_001636 (group 3), covering the blood group type gene *RHCE* (1p36.11), and a loss in patients P_000557 (group 2) and P_000515 (group 3) of which only the *ETFDH* gene is affected in both patients (4q32.1). None of the CNVs present in any of our patients affected known HSCR genes. The DECIPHER (https://decipher.sanger.ac.uk/) contains 18 HSCR patients with associated anomalies with one or more CNVs. Of these, 10 are *de novo*. The 22q11 deletion described in one of these patients (ID 249397) had overlap with the 22q11 deletion found in patient P_000561 (chr22:21032298-21630630). Several genes are located in this region, but only *LZTR1* is overexpressed in the ENS. However, this gene is loss of function tolerant (PLI=0). One of the losses described in other DECIPHER patient (ID 249405) overlaps with the 1q44 loss present in patient P_002431, with *AKT3* as the only gene affected by both CNVs. This gene is loss of function intolerant (PLI=1)

#### Ove*rlapping CNV with a published HSCR cohort*

Only five genes in a CN loss in our cohort were also affected in a previously described HSCR cohort^1^. However, none of them was dosage sensitive, and only Arhgap22 was overexpressed in the developing mouse ENS. The expression of Aif1 was highly upregulated in the developing mouse intestine compared to the ENS, and we could not measure differential expression of three genes (Apom, Galnt11, and Csnk2b). Unfortunately, as only micro-array based expression data of the ENS was available, we cannot reliably distinguish between not expressed and not differentially expressed genes between the ENS and the intestine. However, CSNK2B was highly expressed in human embryonic colon (EW12-EW16).

### S4: Statistical comparisons

|  | *_group 1 vs group 2_* | *_group 1 vs group 3_* | *_group 2 vs group 3_* | *_Group 1 vs group 4_* | *_Group 2 vs group 4_* | *_Group 3 vs group 4_* |  | *_group 1 (n=23)_* | *_group 2 (n=15)_* | *_group 3 (n=20)_* | *_group 4 (n=326)_* | *_group 5 (n=727)_* |
| --- | --- | --- | --- | --- | --- | --- | --- | --- | --- | --- | --- | --- |
| _rare CNV Size_ | *_5.576E-02_* | *_3.908E-02_* | *_4.680E-01_* | *_3.250E-06_* | *_4.615E-01_* | *_6.471E-01_* |  | *_708952_* | *_69472_* | *_109500_* | *_156518_* | *_ND_* |
| _rare Loss Size_ | *_1.001E-01_* | *_5.913E-02_* | *_8.377E-01_* | *_3.641E-07_* | *_7.049E-01_* | *_6.996E-01_* |  | *_574906_* | *_20779_* | *_24817_* | *_55062_* | *_ND_* |
| _ENS genes in a rare CNV_ | *_8.166E-02_* | *_6.514E-02_* | *_5.257E-01_* | *_3.148E-08_* | *_5.513E-01_* | *_9.468E-01_* |  | *_3.217E+00_* | *_3.333E-01_* | *_5.500E-01_* | *_5.736E-01_* | *_ND_* |
| _ENS genes in a rare Loss_ | *_1.010E-01_* | *_6.637E-02_* | *_5.536E-01_* | *_1.760E-10_* | *_5.326E-01_* | *_7.480E-01_* |  | *_2.783E+00_* | *_6.667E-02_* | *_1.500E-01_* | *_2.178E-01_* | *_ND_* |
| _Variant intolerant gene in a rare CNV_ | *_2.018E-01_* | *_8.578E-02_* | *_3.776E-01_* | *_3.359E-05_* | *_9.782E-01_* | *_3.975E-01_* |  | *_1.522E+00_* | *_3.333E-01_* | *_1.500E-01_* | *_3.405E-01_* | *_ND_* |
| _Variant intolerant gene in a rare loss_ | *_2.177E-01_* | *_1.058E-01_* | *_2.833E-01_* | *_1.384E-06_* | *_7.735E-01_* | *_5.715E-01_* |  | *_1.348E+00_* | *_2.000E-01_* | *_5.000E-02_* | *_1.442E-01_* | *_ND_* |
| _ENS and variant intolerant gene in a rare Loss_ | *_2.551E-01_* | *_1.595E-01_* | *_8.394E-01_* | *_3.915E-04_* | *_8.398E-01_* | *_9.720E-01_* |  | *_3.043E-01_* | *_6.667E-02_* | *_9.725E-01_* | *_5.215E-02_* | *_ND_* |
| _RSnc with Sanger sequenced cases and with HCS850k genotyped controls_ | _7.529E-02_ | _1.428E-01_ | _1.526E-03_ | _8.990E-16_ | _2.556E-04_ | _2.290E-22_ |  | _4.420E+00_ | _3.587E+00_ | _4.919E+00_ | _2.619E+00_ | *_ND_* |
| _RSnc with Sanger sequenced cases and with GSA genotyped controls_ | _4.027E-02_ | _2.077E-02_ | _2.293E-04_ | _4.598E-14_ | _9.606E-04_ | _1.219E-23_ |  | _4.707E+00_ | _3.697E+00_ | _5.660E+00_ | *_ND_* | _2.545E+00_ |
| _RSnc with GSA genotyped cases and controls_ | _1.369E-01_ | _3.544E-01_ | _1.267E-02_ | _4.056E-08_ | _4.942E-05_ | _9.496E-20_ |  | _4.885E+00_ | _4.061E+00_ | _5.363E+00_ | *_ND_* | _2.545E+00_ |

*The number and size of rare CNVs, the number of rare losses and gains, the number of genes intolerant to variation (SNVs and CNVs), the number of genes overexpressed in mouse ENS per rare CNV, and the relative weighted risk score, were determined and compared for the different groups with a single ANOVA test. If group differences existed (P<0.05), we determined which subgroups were significantly different, using a two-tailed T-test Abbreviations: CNV; Copy Number Variation, ENS; Enteric Nervous System, NA; not available. Higher values in green, lower values red. Two-tailed p-values.*

**S4 (continued): Statistical comparisons**
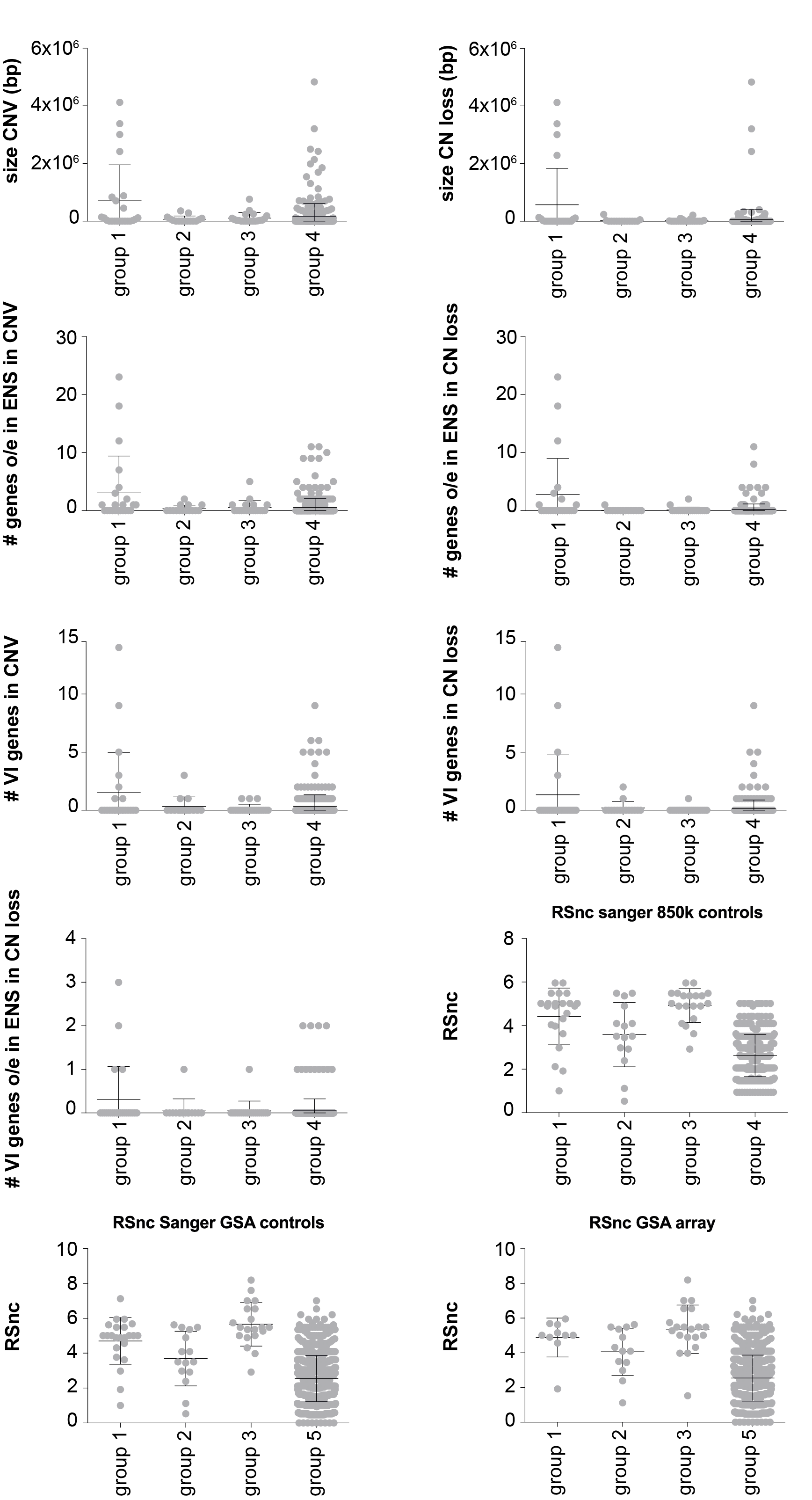

**S4 (continued): Statistical comparisons**

#### HSCR patients with associated anomalies without a known mutation, have larger rare CNVs.

The number of rare CNVs (P=0.385), number of rare losses (P= 0.420) and number of rare gains (P= 0.731), did not differ between unaffected individuals and any of the HSCR groups. Absence of a rare CNV per patient also did not differ between any of the groups (P= 0.363). Next, we compared the size of rare CN gains and of rare losses between groups. As described previously, rare large losses were enriched in HSCR patients with associated anomalies^1^. If CN losses contain dosage sensitive genes overexpressed in the developing ENS, this could suggest that the CNVs are related to HSCR development in these patients. Comparing CNV sizes showed a significant difference between unaffected individuals (Group 4), and HSCR-AAM without a known mutation (Group 1). The size of the loss and not gain of the CNV is responsible for this difference. This effect was not present in Groups 2 or 3. We find that loss size rather than number is associated with HSCR development. However, a previous study described that HSCR patients with associated anomalies have more CNVs compared to isolated HSCR patients and unaffected controls^1^. An explanation for this discrepancy might be that we used a CNV frequency cut-off of 0.026%, to HSCR prevalence (0.02-0.03%). These CNVs are too rare to find a significant association in our study cohort size.

#### CNVs with genes overexpressed in the ENS and intolerant to variation and/or CN changes

In total, 1216 genes or transcripts were present in group 1-3 and group 4. Of these, 514 did not have a known mouse orthologue or probes in microarray datasets (n=279 in group 1-3, n=235 in group 4). There was no differential expression of 472 genes (n=179 in group 1-3, n=293 in group 4). A total of 230 genes affected by a rare CNV, had upregulated expression in the developing mouse ENS (n=91 in group 1-3, n=139 in group 4). We considered these genes to be important for ENS development. Rare CNVs present in HSCR patients contain these ENS genes: 74 in group 1 CNVs, versus 5 in group 2, 11 in group 3 and 187 in group 4 (P= 4.565E^-6^). This result is mostly dependent on the overrepresentation of ENS genes in rare losses in complex-HSCR patients with associated anomalies and without a known mutation: 68 in group 1 CNVs, versus 1 in group 2, 3 in group 3 and 71 in group 4 (P= 4.564E^-6^) (see supplementary data 5). Next, we evaluated the number of ENS genes intolerant to variation and found no statistically significant differences: 10 in CNVs of group 1, versus 3 in group 2, 2 in group 3 and 51 in group 4 (P= 0.093). The CN losses did differ significantly between groups, as the majority of ENS genes intolerant to variation were located in losses in group 1 (n=8), versus 1 in group 2, 2 in group 3 and 22 in group 4 (P= 0.014). All rare CNVs with genes overexpressed in the mouse ENS, are depicted in S7. Comparing the gene content of the rare CNVs between unaffected controls (Group 4) and HSCR subgroups 1 to 3, revealed that the rare CNVs from Group 1 contained more of these intolerant genes (P=0.0002). Moreover, the CN losses in this group contained more intolerant genes overexpressed in the ENS (P=0.001), compared to Group 4.

### S5: The Z-score distributions of total CNV size, CNV loss and gain of controls and HSCR patients.

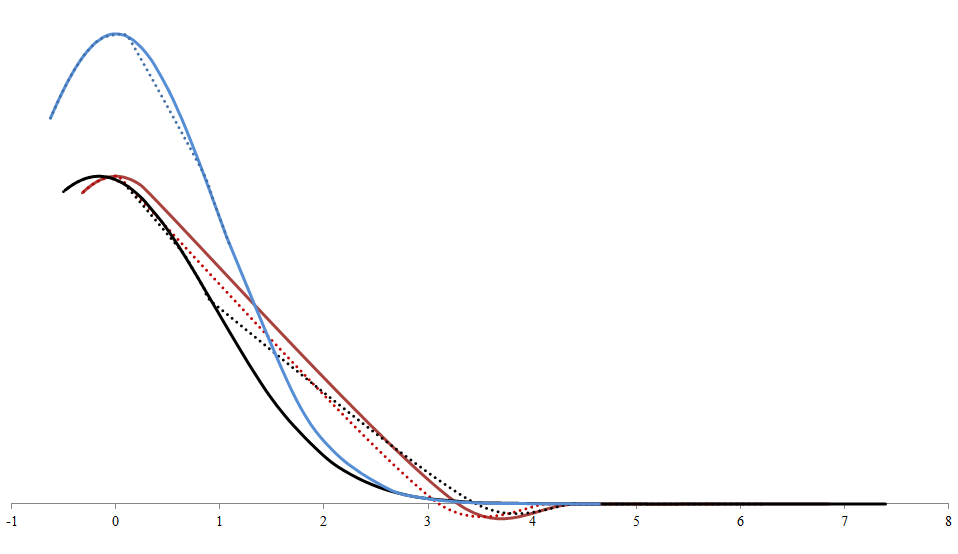
 *Depicted are the Z-score distributions of the all rare CNV (black lines), rare Copy Number Losses (red lines) and Gains (blue lines) Controls in continuous lines, all HSCR patients in dotted lines. Limited negative z-scores due to the size limit cut-off of 20kb. The z-score distributions of CNV sizes were comparable between groups strongly suggesting that the outliers are responsible for the statistical significant groupwise-differences.*

### S6: HSCR disease gene characteristics

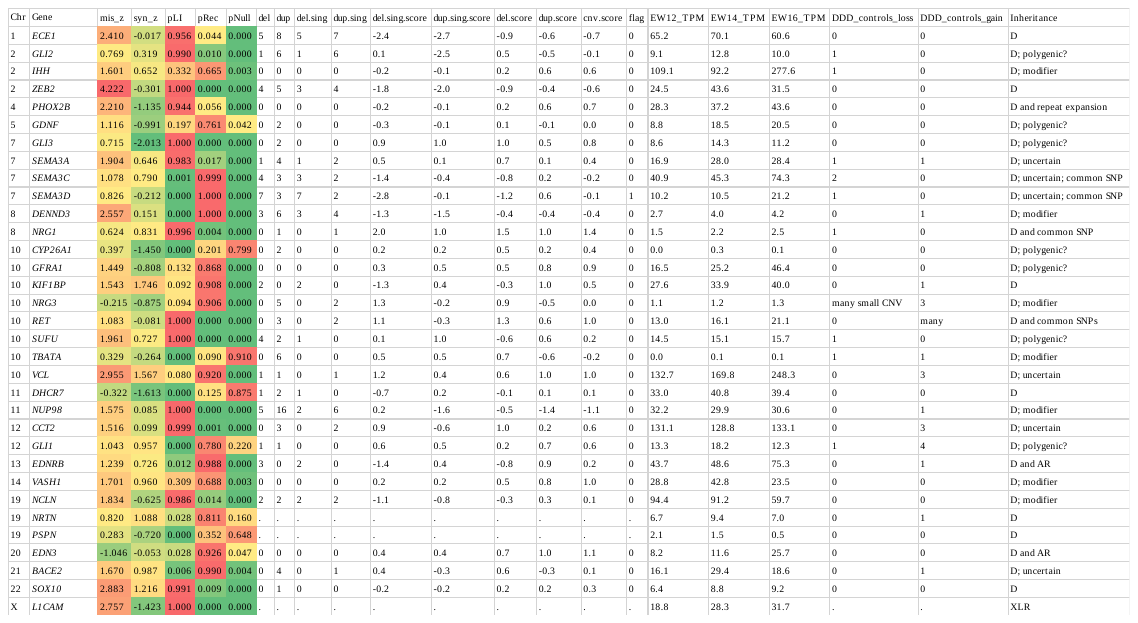

*Most known HSCR genes are intolerant to genetic variation and are rarely impacted by CNVs in unaffected individuals^2; 3^. These genes have been described to be impacted by CNVs in HSCR patients^4; 5^, although this does not seem to be a frequent phenomenon, as in our cohort we did not detect any CNV impacting a known HSCR gene.*

### S7: Rare CNV with genes overexpressed in mouse ENS between E11.5 and E15.5

| **P-number** | **Chromosome Region** | **Event** | **Length** | **Cytoband** | **Probes** | **Class** | **Sex** | **ENS gene(s)** |
| --- | --- | --- | --- | --- | --- | --- | --- | --- |
| **P_000302** | chr3:14,406,477-14,509,088 | CN Gain | 102612 | p25.1 | 53 | LD | F | **SLC6A6** |
| **P_000479** | chr12:9,245,492-9,308,543 | CN Gain | 63052 | p13.31 | 24 | VUS | M | A2M |
| **P_000479** | chr2:40,624,267-40,646,501 | CN Loss | 22235 | p22.1 | 11 | LD | M | **SLC8A1** |
| **P_000494** | chr12:128,208,742-128,917,555 | CN Gain | 708814 | q24.32 | 96 | VUS | M | TMEM132C |
| **P_000498** | chr1:152,286,216-152,323,703 | CN Gain | 37488 | q21.3 | 11 | VUS | F | FLG |
| **P_000502** | chr15:80,527,215-80,603,142 | CN Gain | 75928 | q25.1 | 22 | VUS | F | CTXND1 |
| **P_000512** | chr6:28,005,012-31,683,185 | CN Loss | 3678174 | p22.1 - p21.33 | 403 | LD | F | 6M1-18, ABHD16A, ATAT1, ATP6V1G2, DDR1, DPCR1, FLOT1, **GABBR1**, **GNL1**, HLA-H, IER3, MUCL3, OR11A1, OR2J2, OR2J3, PGBD1, PPP1R11, PP1R18, **TUBB**, ZKSCAN4, ZNRD1-AS1, ZNRD1ASP, ZSCAN31 |
| **P_000520** | chr18:45,755,986-45,787,673 | CN Gain | 31688 | q21.1 | 13 | VUS | M | ZBTB7C |
| **P_000537** | chr10:49,033,586-52,417,694 | CN Loss | 3384109 | q11.22 - q11.23 | 183 | LD | M | ARHGAP22, C10orf128, CHAT, FAM21A, **MAPK8**, NCOA4, SLC18A3, TIMM23, TIMM23B, TMEM273, VSTM4, WASHC2A |
| **P_000540** | chrX:154,277,428-154,299,482 | Hemizygous | 22055 | q28 | 5 | VUS | M | CMC4, FUNDC2, MTCP2 |
| **P_000557** | chr11:62,251,301-62,298,871 | CN Gain | 47571 | q12.3 | 25 | LD | M | **AHNAK** |
| **P_000561** | chr22:18,861,209-21,630,630 | CN Loss | 2769422 | q11.21 | 446 | LD | M | ARVCF, BCRP2, C22orf29, C22orf39, CDC45, COMT, DGCR14, DGCR2, ESS2, GP1BB, LZTR1, RIMBP3, RTL10, SLC7A4, UFD1, **UFD1L**, ZDHHC8, ZNF74 |
| **P_000567** | chr17:58,076,721-60,362,868 | CN Loss | 2286148 | q23.1 - q23.2 | 74 | LD | M | BCAS3, HEATR6, **TBX2**, **USP32** |
| **P_000573** | chr2:216,214,577-216,299,733 | CN Loss | 85157 | q35 | 49 | VUS | F | FN1 |
| **P_000579** | chr3:60,468,409-60,490,104 | CN Loss | 21696 | p14.2 | 16 | LD | M | **FHIT** |
| **P_000582** | chr7:4,929,022-5,218,030 | CN Gain | 289009 | p22.1 | 59 | VUS | M | MMD2 |
| **P_000582** | chr2:102,658,576-102,847,088 | CN Gain | 188513 | q11.2 - q12.1 | 57 | VUS | M | IL1R1, IL1RL2 |
| **P_000582** | chr7:5,239,584-5,401,976 | CN Gain | 162393 | p22.1 | 54 | VUS | M | SLC29A4, WIPI2 |
| **P_001632** | chr22:18,687,210-19,060,954 | CN Gain | 373745 | q11.21 | 75 | VUS | M | DGCR2; known modifier CNV |
| **P_001637** | chr2:10,664,398-10,914,786 | CN Gain | 250389 | p25.1 | 70 | VUS | M | NOL10 |
| **P_001763** | chr18:65,699,090-66,534,856 | CN Gain | 835767 | q22.1 | 228 | VUS | F | TMX3 |
| **P_002431** | chr1:243,963,527-244,016,804 | CN Loss | 53278 | q44 | 9 | LD | F | **AKT3** |
| **P_002431** | chr7:95,845,896-96,004,178 | CN Loss | 158283 | q21.3 | 16 | VUS | F | SLC25A13 |
| **P_002450** | chr9:28,393,380-28,462,962 | CN Loss | 69583 | p21.1 | 21 | VUS | M | LINGO2 |
| **P_002455** | chr22:21,802,791-22,555,544 | CN Gain | 752756 | q11.21 - q11.22 | 169 | VUS | M | CCDC116, **MAPK1**, PPM1F, SDF2L1, TMEM191C, **YDJC**, YPEL1 |
| **P_002459** | chr12:94,767,704-94,880,489 | CN Loss | 112786 | q22 | 27 | VUS | M | CCDC41, CEP83 |
| Abbreviations: CN; Copy Number, LD; Likely deleterious, VUS; variant of unknown significance, M; Male, F, Female. In yellow/bold overexpressed in ENS and intolerant to variation. Chromosomal regions according to build hg19. | | | | | | | | |

### S8: table 1 patient discussion

Please contact the corresponding author for access to the data presented in S8.

### S9: Variant prioritization and WGS variant burden test results

### Variant prioritization in loss of function genes (Figure 1a)

To determine whether a gene affected by a rare putative deleterious CNV was predicted to be constraint (intolerant to genetic variation)^42; 43^ we used a threshold of probability of Loss of function Intolerant (pLI > 0.85), Probability of recessive (Prec 0.90), synonymous or missense z-score of at least 3 for missense variants and >1 for deletions and duplications.We used capture-specific controls to eliminate technical noise in (1) a WES cohort of sporadic HSCR (n=76, 149 controls) and (2) a Whole Genome Sequencing (WGS) cohort of 443 short segment HSCR patients and 493 unaffected controls^44^. Variants from WES data previously generated, were prioritized as follows: an allele frequency below 1% in *in-house* unaffected controls (n=906); affect a constraint gene and have an allele frequency of maximum 0.01 for homozygous recessive variants and of 0.001 for heterozygous variants, in GnomAD. We used a CADD score of 20 as a measure for deleteriousness for missense variants. All variants within two bases of an intron-exon boundary were considered to affect splicing, and were included in the “loss of function” category when considering gene constraint. Using RVTESTS^45^, a variant burden test was done comparing the variant burden in 443 short segment HSCR patients and 493 controls (WGS)^44^ using an allele frequency of maximum 0.01 for homozygous recessive variants and of 0.001 for heterozygous variants, in GnomAD. We used a CADD score of 20 as a measure for deleteriousness for missense variants. All rare putative deleterious loss of function variants unique to the HSCR cohort in constraint genes are described in table 1 and were uploaded to the ClinVar database (<https://www.ncbi.nlm.nih.gov/clinvar/>).

Abbreviations: NVAR: number of variants, NCASEHET: number of heterozygous variants in cases, NCTRLHET: number of heterozygous variants in controls

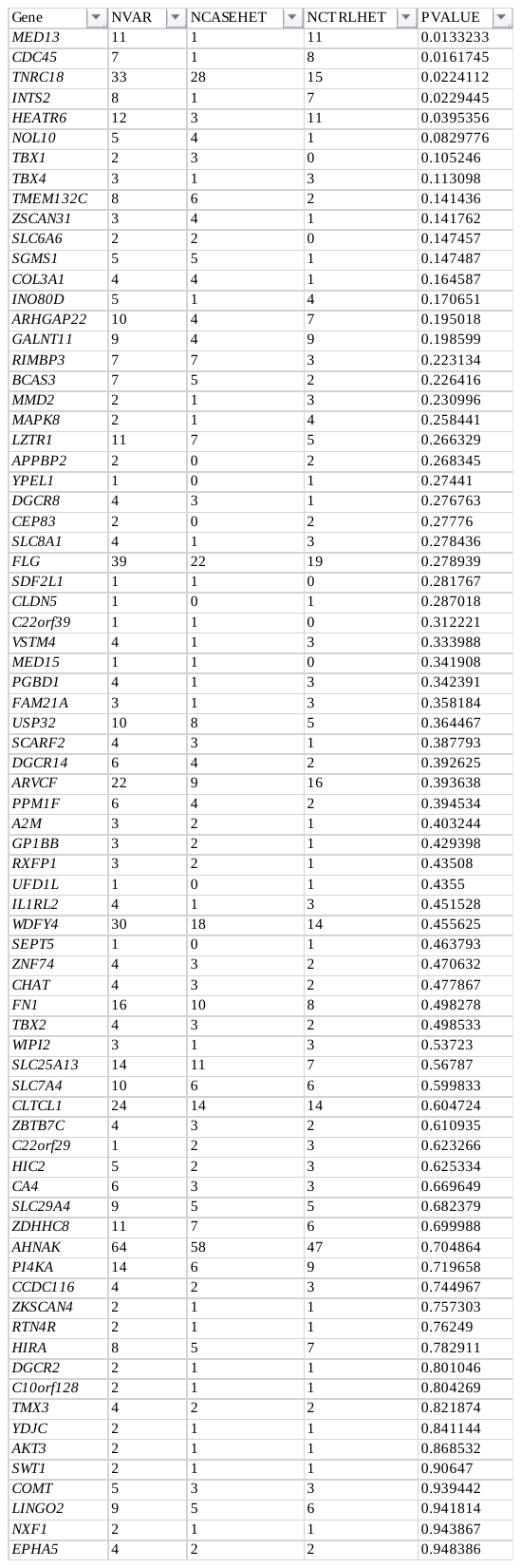

### S10 Risk haplotype markers and odds ratio’s used to calculate noncoding risk scores

Sanger sequencing was used to genotype all patients for SNPs known to be associated to HSCR^46-49^. Primer sequences can be found in S11. We used (proxy) SNPs present on the GSAMD-v1 chip to determine the Rotterdam population background for these risk haplotypes (n= 388 females, 339 males), as well as the three HSCR subgroups from the SNP-array platform (S10). The relative weighted risk score of published common and relatively rare risk alleles near *RET* (rs2506030, rs7069590, rs2435357), *NRG1* (rs7005606) and *SEMA3C/D* (rs11766001, rs80227144), was calculated using the formula below^47-50^.

$\left( \left( LnOR risk allele 1 \right)*allele count \right)+(\left( LnOR risk allele 2 \right)*allele count)+ etc.$

We also determined the risk scores of the CNV control samples (n=326). However, we could only do this for the *RET* and *NRG1*, as no suitable proxy SNPs were present on the HCS 850k platform for the SEMA3C/D haplotypes.

| Gene | risk/non-risk allele | Risk allele frequency | Odds ratio (95% CI) | P | OR | GSAMD v1 | risk/non-risk | D' | R^2^ | Ref |
| --- | --- | --- | --- | --- | --- | --- | --- | --- | --- | --- |
| *SEMA3C/D* | **rs11766001:C/A** | **0.22/0.15** | **1.6 (1.3–2.0)** | **1.0 × 10−4** | **1.6** | **rs4374933** | **T/C** | **0.9834** | **0.6221** | **^47^** |
| *SEMA3C/D* | **rs80227144: C/A** | **0.14/0.03** | **5.2 (3.09–8.73)** | **4.0×10-4** | **5.2** | **rs80227144** | **C/A** | **1** | **1** | **^47^** |
| *RET^#^* | **rs2506030: G/A** | **0.56/0.41** | **1.8 (1.5-2.2)** | **4.46×10-11** | **1.8** | **GSA-rs2506030** | **G/A** | **1** | **1** | **^49^** |
| *RET^#^* | **rs7069590: T/C** | **0.84/0.76** | **1.7 (1.4-2.2)** | **4.36×10-6** | **1.7** | **rs71505660** | **C/A** | **0.9946** | **0.984** | **^49^** |
| *RET^#^* | **rs2435357: T/C** | **0.58/0.25** | **4.01 (3.33–4.84)** | **2.98×10-48** | **4.01** | **rs2435344** | **T/C** | **0.9841** | **0.9338** | **^49^** |
| *RET*^@^ | rs9282834: A/G | 0.05/0.03 | 1.80 (1.06–3.04) | 0.029 | 1.8 | NA (to rare) | NA | NA | NA | ^50^ |
| *RET* | rs2505998: A/C | 0.64/0.22 | 4.17 (3.23–5.26) | 1.1×10-28 | 4.17 | rs2435344 | C/A | 0.9838 | 0.9088 | ^50^ |
| *NRG1* | **rs7005606: T/G** | **0.54/0.42** | **1.64 (1.25–2.15)** | **4.7×10-10** | **1.64** | **rs4733130** | **T/C** | **1** | **1** | **^50^** |
| Gene | **Risk haplotype** | **Risk haplotype frequency** | **Odds ratio (95% CI)** | **P** | **OR** |  |  |  |  | **Ref** |
| *RET* | ATT# | 0.14/0.08 | 3.13 (2.17-4.50) | 8.31 × 10−10 | - |  |  |  |  | ^49^ |
| *RET* | GTT# | 0.42/0.16 | 4.40 (3.26-5.94) | 3.62 × 10−22 | - |  |  |  |  | ^49^ |
| *RET* | TA^@^ | 0.05/0.03 | 20.3 (9.31–44.4) | 2.98×10-48 | - |  |  |  |  | ^50^ |

*Highlighted SNPS* ***in grey/ bold*** *are the risk alleles and Odds ratio’s used in the polygenic risk score calculation. ^#^We did not account for the increased risk of having the two main risk haplotype combinations[1] ^@^ this risk haplotype was not present in this patient cohort. Sanger sequencing was used to genotype all patients for SNPs known to be associated to HSCR^46-49^., Patient and controls genotypes were determined using SNPs from the GSAMD-v1 platform. D’and R2 derived from (*[*https://ldlink.nci.nih.gov/*](https://ldlink.nci.nih.gov/)*) in European population. Primer sequences can be found in supplementary data S12.*

### S11 description of individual patient genetic risk profiles

| **patien** | **segment** | **Sex** | **group** | **rs2506030** | **rs7069590** | **rs2435357** | **rs80227144** | **rs7005606** | **RSnc** | **RET nc risk^$^** | **ENS Gain** | **ENS loss** | **Zebrafish** | **Variant(s) found in indep. cohort** |
| --- | --- | --- | --- | --- | --- | --- | --- | --- | --- | --- | --- | --- | --- | --- |
| **P_000572** | TIA | Male | 1 | 0 | 1 | 0 | 0 | 1 | 1,00 | - | 0 | 0 | 0 | 0 |
| **P_000520** | Short | Male | 1 | 0 | 1 | 1 | 0 | 0 | 1,92 | ATT | 1 | 0 | 0 | 0 |
| **P_000540** | Short | Male | 1 | 1 | 1 | 1 | 0 | 1 | 2,98 | GTT or ATT | 0 | 1 | 0 | 0 |
| **P_000562** | Short | Male | 1 | 2 | 2 | 1 | 0 | 0 | 3,63 | GTT | 0 | 0 | 0 | 0 |
| **P_002459** | Short | Male | 1 | 1 | 2 | 0 | 1 | 1 | 3,77 | - | 0 | 1 | 0 | 0 |
| **P_000528** | Short | Male | 1 | 0 | 2 | 2 | 0 | 1 | 4,31 | ATT and ATT | 0 | 0 | 0 | 0 |
| **P_000537** | Short | Male | 1 | 2 | 2 | 1 | 0 | 2 | 4,57 | GTT | 0 | 1 | 1 | 3 |
| **P_000478** | Short | Male | 1 | 1 | 2 | 2 | 0 | 1 | 4,90 | ATT and GTT | 0 | 0 | 0 | 0 |
| **P_000561** | Short | Male | 1 | 1 | 2 | 2 | 0 | 1 | 4,90 | ATT and GTT | 0 | 1 | 1 | 1 |
| **P_001763** | Short | Female | 1 | 1 | 2 | 2 | 0 | 1 | 4,90 | ATT and GTT | 1 | 0 | 0 | 0 |
| **P_000482** | Short | Male | 1 | 2 | 2 | 2 | 0 | 0 | 5,01 | GTT and GTT | 0 | 0 | 0 | 0 |
| **P_000536** | abnormal | Female | 1 | 2 | 2 | 2 | 0 | 0 | 5,01 | GTT and GTT | 0 | 0 | 0 | 0 |
| **P_000555** | Short | Female | 1 | 2 | 2 | 2 | 0 | 0 | 5,01 | GTT and GTT | 0 | 0 | 0 | 1 |
| **P_000568** | Short | Male | 1 | 2 | 2 | 2 | 0 | 0 | 5,01 | GTT and GTT | 0 | 0 | 0 | 0 |
| **P_000573** | Short | Female | 1 | 2 | 2 | 2 | 0 | 0 | 5,01 | GTT and GTT | 0 | 1 | 0 | 0 |
| **P_002450** | Long | Male | 1 | 2 | 2 | 2 | 0 | 0 | 5,01 | GTT and GTT | 0 | 1 | 0 | 1 |
| **P_000553** | Short | Male | 1 | 2 | 2 | 2 | 0 | 1 | 5,48 | GTT and GTT | 0 | 0 | 0 | 0 |
| **P_002343** | Short | Male | 1 | 2 | 2 | 2 | 0 | 1 | 5,48 | GTT and GTT | 0 | 0 | 0 | 0 |
| **P_000567** | Short | Male | 1 | 1 | 2 | 1 | 1 | 2 | 5,63 | GTT or ATT | 0 | 1 | 1 | 3 |
| **P_002455** | Short | Male | 1 | 2 | 1 | 1 | 1 | 2 | 5,68 | GTT or ATT | 1 | 0 | 0 | 0 |
| **P_000494** | Short | Male | 1 | 2 | 2 | 2 | 0 | 2 | 5,95 | GTT and GTT | 1 | 0 | 0 | 0 |
| **P_000512** | Short | Female | 1 | 2 | 2 | 2 | 0 | 2 | 5,95 | GTT and GTT | 0 | 1 | 1 | 2 |
| **P_000559** | TCA | Male | 1 | 2 | 2 | 2 | 1 | 1 | 7,13 | GTT and GTT | 0 | 0 | 0 | 0 |
| **P_000302** | Short | Female | 2 | 0 | 1 | 0 | 0 | 0 | 0,53 | - | 1 | 0 | 0 | 1 |
| **P_002442** | Long | Male | 2 | 1 | 1 | 0 | 0 | 0 | 1,12 | - | 0 | 0 | 0 | 0 |
| **P_004502** | Short | Male | 2 | 0 | 1 | 1 | 0 | 1 | 2,39 | ATT | 0 | 0 | 0 | 0 |
| **P_000557** | TCA | Male | 2 | 0 | 2 | 1 | 0 | 1 | 2,92 | ATT | 1 | 0 | 0 | 0 |
| **P_000479** | Long | Male | 2 | 1 | 1 | 1 | 0 | 1 | 2,98 | GTT or ATT | 1 | 1 | 1 | 0 |
| **P_000518** | Short | Female | 2 | 1 | 1 | 1 | 0 | 2 | 3,45 | GTT or ATT | 0 | 0 | 0 | 0 |
| **P_000534** | Short | Female | 2 | 1 | 2 | 1 | 0 | 1 | 3,51 | GTT or ATT | 0 | 0 | 0 | 0 |
| **P_000570** | Short | Male | 2 | 1 | 2 | 1 | 0 | 1 | 3,51 | GTT or ATT | 0 | 0 | 0 | 0 |
| **P_000502** | Short | Female | 2 | 2 | 2 | 1 | 0 | 1 | 4,10 | GTT | 1 | 0 | 0 | 0 |
| **P_000526** | Short | Female | 2 | 2 | 2 | 1 | 0 | 1 | 4,10 | GTT | 0 | 0 | 0 | 0 |
| **P_000576** | Short | Female | 2 | 1 | 2 | 2 | 0 | 1 | 4,90 | ATT and GTT | 0 | 0 | 0 | 0 |
| **P_000566** | Short | Male | 2 | 1 | 2 | 2 | 0 | 2 | 5,37 | ATT and GTT | 0 | 0 | 0 | 0 |
| **P_000480** | Short | Male | 2 | 2 | 2 | 2 | 0 | 1 | 5,48 | GTT and GTT | 0 | 0 | 0 | 0 |
| **P_000486** | TCA | Female | 2 | 2 | 2 | 2 | 0 | 1 | 5,48 | GTT and GTT | 0 | 0 | 0 | 0 |
| **P_000544** | Long | Male | 2 | 1 | 2 | 1 | 1 | 2 | 5,63 | GTT or ATT | 0 | 0 | 0 | 0 |
| **P_000577** | Short | Male | 3 | 0 | 2 | 1 | 0 | 1 | 2,92 | ATT | 0 | 0 | 0 | 0 |
| **P_000515** | Short | Male | 3 | 1 | 2 | 1 | 0 | 2 | 3,98 | GTT or ATT | 0 | 0 | 0 | 0 |
| **P_001638** | Short | Male | 3 | 0 | 2 | 2 | 0 | 1 | 4,31 | ATT and ATT | 0 | 0 | 0 | 0 |
| **P_000578** | Long | Male | 3 | 1 | 2 | 2 | 0 | 1 | 4,90 | ATT and GTT | 0 | 0 | 0 | 0 |
| **P_000579** | Short | Male | 3 | 2 | 2 | 2 | 0 | 0 | 5,01 | GTT and GTT | 0 | 1 | 0 | 0 |
| **P_001635** | Short | Female | 3 | 2 | 2 | 2 | 0 | 0 | 5,01 | GTT and GTT | 0 | 0 | 0 | 0 |
| **P_000490** | Short | Female | 3 | 2 | 2 | 1 | 1 | 0 | 5,27 | GTT | 0 | 0 | 0 | 0 |
| **P_000498** | Long | Female | 3 | 1 | 2 | 2 | 0 | 2 | 5,37 | ATT and GTT | 1 | 0 | 0 | 0 |
| **P_000554** | Short | Male | 3 | 1 | 2 | 2 | 0 | 2 | 5,37 | ATT and GTT | 0 | 0 | 0 | 0 |
| **P_000505** | Short | Male | 3 | 2 | 2 | 2 | 0 | 1 | 5,48 | GTT and GTT | 0 | 0 | 0 | 0 |
| **P_000514** | Short | Male | 3 | 2 | 2 | 2 | 0 | 1 | 5,48 | GTT and GTT | 0 | 0 | 0 | 0 |
| **P_001639** | Short | Male | 3 | 2 | 2 | 2 | 0 | 1 | 5,48 | GTT and GTT | 0 | 0 | 0 | 0 |
| **P_000582** | Short | Male | 3 | 2 | 2 | 1 | 1 | 1 | 5,74 | GTT | 3 | 0 | 0 | 0 |
| **P_001636** | Short | Male | 3 | 2 | 2 | 2 | 0 | 2 | 5,95 | GTT and GTT | 0 | 0 | 0 | 0 |
| **P_000575** | Short | Male | 3 | 1 | 2 | 2 | 1 | 1 | 6,55 | ATT and GTT | 0 | 0 | 0 | 0 |
| **P_001632** | Short | Male | 3 | 1 | 2 | 2 | 1 | 1 | 6,55 | ATT and GTT | 1 | 0 | 0 | 0 |
| **P_000552** | Short | Male | 3 | 1 | 2 | 2 | 1 | 2 | 7,02 | ATT and GTT | 0 | 0 | 0 | 0 |
| **P_002431** | Short | Female | 3 | 1 | 2 | 2 | 1 | 2 | 7,02 | ATT and GTT | 0 | 2 | 0 | 0 |
| **P_000450** | Long | Male | 3 | 2 | 2 | 2 | 1 | 2 | 7,60 | GTT and GTT | 0 | 0 | 0 | 0 |
| **P_001637** | Short | Male | 3 | 1 | 2 | 2 | 2 | 1 | 8,19 | ATT and GTT | 1 | 0 | 0 | 0 |

*Depicted are the individual patients characteristics, number of predisposing haplotypes (as represented by the risk SNPs) and the noncoding risk score (RSnc). Additionally, the RET risk haplotypes as described in S10 are depicted. In blue the number of ENS overexpressed genes in a gain, in red the number of ENS overexpressed genes in a loss. Depicted in blue: (1) functional evidence from zebrafish studies and (2) the number additional patients containing putative deleterious variants in genes within the rare CNVs . ^$^No phasing was performed to discern the most likely haplotype*

### S12: primer sequences common predisposing SNPs

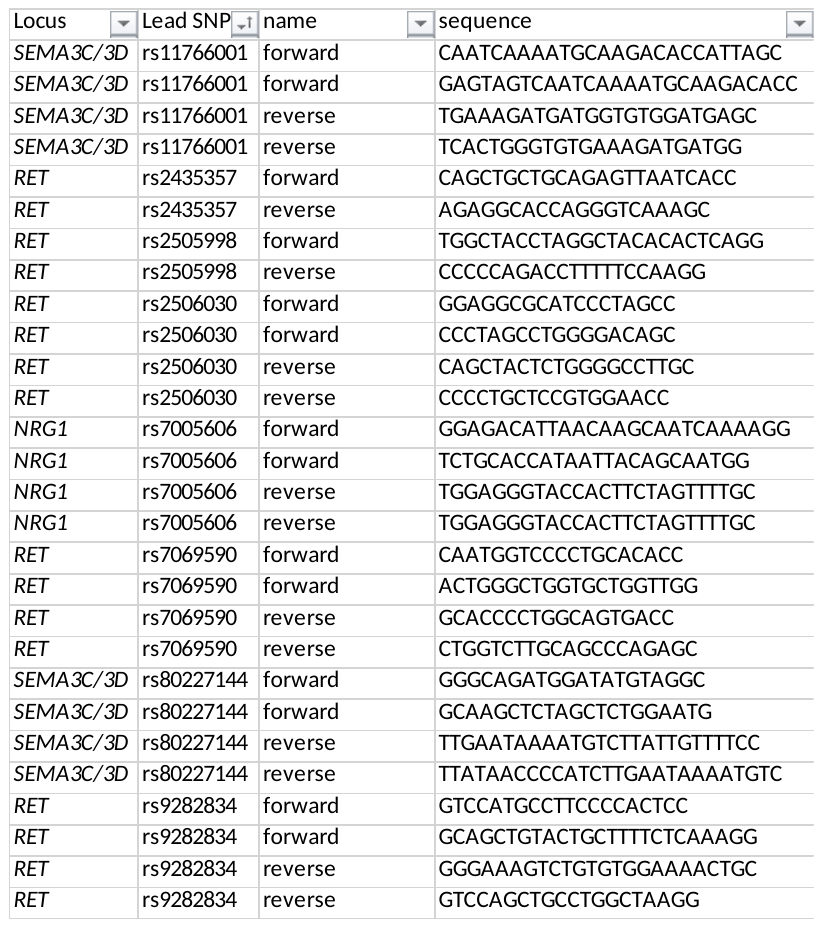

### S13: Details of the gRNA sequences used

| Gene | Position | Strand | Sequence | PAM | On-target score | Off-target score |
| --- | --- | --- | --- | --- | --- | --- |
| *tubb1* | 49887677 | - | TGGTTGAGATCTCCATAGGT | TGG | 71 | [92](https://eu.idtdna.com/site/order/designtool/index/CRISPR_CUSTOM) |
| *gnl1* | 21665680 | + | TCGGCTGCTGTGACCTGACC | CGG | 62 | [95](https://eu.idtdna.com/site/order/designtool/index/CRISPR_CUSTOM) |
| *tbx2a* | 56513198 | - | TTTCAAGGGTCTCGAGCCAG | AGG | 77 | [95](https://eu.idtdna.com/site/order/designtool/index/CRISPR_PREDESIGN) |
| *tbx2b* | 27365003 | - | GGTGGTGACTCAAAGCCGGA | TGG | 65 | [94](https://eu.idtdna.com/site/order/designtool/index/CRISPR_PREDESIGN) |
| *ufd1l* | 4768775 | - | GACCGCCCTCTTCCAACAAC | AGG | 56 | [96](https://eu.idtdna.com/site/order/designtool/index/CRISPR_CUSTOM) |
| *usp32* | custom | - | CAGACGTTTGAGCTCCACAT | CGG | 70 | [82](https://eu.idtdna.com/site/order/designtool/index/CRISPR_CUSTOM) |
| *akt3a* | 11233467 | - | CGGATACAAGGAGAAGCCAC | AGG | 96 | [97](https://eu.idtdna.com/site/order/designtool/index/CRISPR_PREDESIGN) |
| *akt3b* | custom | + | GTGAGTACATTAAGAACTGG | AGG | 83 | [83](https://eu.idtdna.com/site/order/designtool/index/CRISPR_PREDESIGN) |
| *gabbr1a* | custom | + | CTGGTTTAAGATCAAAGATC | CGG | 82 | 75 |
| *gabbr1b* | custom | + | GAACACAAGTTCATTCGAGG | GGG | 82 | [89](https://eu.idtdna.com/site/order/designtool/index/CRISPR_PREDESIGN) |
| *slc8a1a* | 30601171 | + | TCCCCAAAGGACGGGTTCAC | CGG | 77 | [97](https://eu.idtdna.com/site/order/designtool/index/CRISPR_PREDESIGN) |
| *slc8a1b* | 22381989 | - | CCAAGCGAAGACCACGCAAA | TGG | 72 | [95](https://eu.idtdna.com/site/order/designtool/index/CRISPR_PREDESIGN) |
| *mapk8a* | 31160246 | + | TAGTAACGGGTCACCACATA | TGG | 85 | [94](https://eu.idtdna.com/site/order/designtool/index/CRISPR_PREDESIGN) |
| *mapk8b* | 2516188 | + | CCACATTTCGATCGAGGACG | TGG | 61 | [98](https://eu.idtdna.com/site/order/designtool/index/CRISPR_PREDESIGN) |

### S14: Primers used for Sanger sequencing

| Primer name | Primer sequence |
| --- | --- |
| tubb1_R | CTTCAGTGTAATGCCCTCGC |
| tubb1_F | AGAGCTCGGTATTGTTGGCT |
| gnl1_F | GCTGGTTGACGAGAGCTTG |
| gnl1_R | TGTTGGGGTCATATCTGCCG |
| tbx2a_F | AACCTTCGTTCTTTCCAGCG |
| tbx2a_R | AGAGGCTTCGATGCTATGTCA |
| tbx2b_F | CCCATGTCAGCTTTTCTCGC |
| tbx2b_R | TTCAATCGCGTAAACACTGC |
| ufd1l_F | TTCAGTGGAAAAGCGTGGTG |
| ufd1l_R | TGTGTCTTCAACTCTATCTGTGT |
| usp32_F | ACTTGAAGAATATCGCACGACT |
| usp32_R | GATGATGATCACGTTGAACTCAC |
| akt3a_F | TGTGCGATTGTGGGTTTGAG |
| akt3a_R | CCAGTAAAAGCAAGTCTCCAGT |
| akt3b_F | TGAACGTCGTGAAAGAGGGA |
| akt3b_R | GGAGTCTGATAATGAGAGCGAC |
| gabbr1a_F | CCGAGGCTTAACCGAGATT |
| gabbr1a_R | TCATGATCCTGTTGTGAAAGTCT |
| gabbr1b_F | CCTTTGGCGTATGATGCAGT |
| gabbr1b_R | GTCCTCGCCTGTGTGAACA |
| slc8a1a_F | TCCTATGAGCTCACGCCAGT |
| slc8a1a_R | TCTCCTGCGTCAAAGTTGTG |
| Slc8a1b_F | AACATCGCAAAGTGAAACACC |
| Slc8a1b_R | CTCCAGTTCCAAAGCCAGAG |
| mapk8a_R | ATCAGAGGGAGCACAAATGG |
| mapk8a_F | CGATTCTTTAGGACCTGAAACC |
| mapk8b_R | TCCATGTCTTACATTTTTGTGGTT |
| mapk8b_F | AATAAAGTGGCCGGTGAGTG |

### S15: gRNA efficiency scores with respective references to the figures

| Gene | Efficiency | R2 | Figure |
| --- | --- | --- | --- |
| *gnl1* | 75% | 0,80 | 2c |
| *ufd1l* | 95% | 0,95 | 2c |
| *usp32* | 85% | 0,87 | 2c |
| *tubb1* | 80% | 0,85 | 2c |
| *akt3a* | 41% | 0,88 | 2c |
| *akt3b* | 67% | 0,77 | 2c |
| *tbx2a* | 86% | 0,86 | 2c |
| *tbx2b* | 68% | 0,91 | 2c |
| *gabbr1a* | 80% | 0,89 | 2c |
| *gabbr1b* | 82% | 0,82 | 2c |
| *slc8a1a* | 84% | 0,9 | 2c |
| *slc8a1b* | 10% | 0,95 | 2c |
| *mapk8a* | 85% | 0,89 | 2c |
| *mapk8b* | 17% | 0,84 | 2c |
| *gnl1* | 96% | 0,96 | 2e |
| *ufd1l* | 96% | 0,96 | 2e |
| *usp32* | 85% | 0,91 | 2e |
| *tubb1* | 85% | 0,86 | 2e |
| *akt3a* | 25% | 0,94 | 2e |
| *akt3b* | 69% | 0,78 | 2e |
| *tbx2a* | 92% | 0,92 | 2e |
| *tbx2b* | 27% | 0,97 | 2e |
| *gabbr1a* | 81% | 0,81 | 2e |
| *gabbr1b* | 89% | 0,9 | 2e |
| *mapk8a* | 87% | 0,89 | 2e |
| *mapk8b* | 29% | 0,88 | 2e |
| *gnl1* | 96% | 0,96 | 2f |

### S16: Statistics and number of fish used per group with respective references to the figures

| Gene | significance | chi-squared | number of larvae | Figure |
| --- | --- | --- | --- | --- |
| *slc8a1* | ***p=0,0073*** | chi-squared = 7,207 | n=15 | 2d |
| *tbx2* | ***p=0,0373*** | chi-squared=4,339 | n=33 | 2d |
| *mapk8* | ***p=0,0022*** | chi-squared=9,346 | n=37 | 2d |
| *gnl1* | p=0,4028 | chi-squared = 0,700 | n=30 | 2d |
| *ufd1l* | ***p=0,0208*** | chi-squared=5,344 | n=40 | 2d |
| *usp32* | NA | NA | n=23 | 2d |
| *tubb1* | NA | NA | n=20 | 2d |
| *akt3* | NA | NA | n=28 | 2d |
| *gabbr1* | P=0,3778 | Chi-squared=0,778 | n=27 | 2d |
| *mapk8* | ***P=0,0107*** | Chi-squared=6,518 | n=20 | 2e |
| *tbx2* | p=0,4073 | Chi-squared=0,687 | n=38 | 2e |
| *gnl1* | p=0,0664 | Chi-squared=3,371 | n=32 | 2e |
| *ufd1l* | p=0,3915 | Chi-squared=0,734 | n=31 | 2e |
| *usp32* | p=0,1847 | Chi-squared=1,759 | n=32 | 2e |
| *tubb1* | p=0,3356 | Chi-squared=0,927 | n=34 | 2e |
| *akt3* | P=0,4157 | Chi-squared=0,663 | n=30 | 2e |
| *gabbr1* | p=0,1481 | Chi-squared=2,091 | n=31 | 2e |
| *gnl1* | ***p=0.0405*** | Chi-squared=4,199 | n=23 | 2f |
